# Epithelial-to-mesenchymal transition lowers the cholesterol pathway, which influences colon tumors differentiation

**DOI:** 10.1101/2021.10.13.21264887

**Authors:** Anaïs Aulas, Maria Lucia Liberatoscioli, Pascal Finetti, Olivier Cabaud, David J. Birnbaum, Daniel Birnbaum, François Bertucci, Emilie Mamessier

## Abstract

Colorectal cancer (CRC) is the second cause of death worldwide. Up to 70% of CRC patients will metastasize. Identify new biomarkers of progression to prevent/delay disease evolution is urgent. Epithelial-to-mesenchymal transition (EMT) is a major program engaged during metastasis. We aim at identifying a combination of new genes that further identify signs of EMT in cancer tissues. We treated HT-29 cells grown in 3D with an EMT Inducing cocktail, and collect them before during and after treatment. For each condition, pan-transcriptomic analyses were done. Genes that were both induced upon EMT induction and inhibited upon EMT release (mesenchymal-to-epithelial transition or MET) were selected. We identified new genes for EMT-MET programs. These genes were used to build a metagene that, when applied to a database of transcriptomic data from primary colorectal tumors (n= 2,239), had an independent prognosis value. Finally, we submitted this metagene to CMap and identified drugs that might affect EMT-MET programs. Statins, well-known inhibitors of cholesterol synthesis, were among them and effectively delayed MET. These data show that cholesterol and EMT pathways are opposite regulators and impact differently tumor differentiation and outcome.

**TRANSLATIONAL RELEVANCE:** Metastases are the main cause of death of colorectal cancer (CRC) and the major issue in CRC management. Understanding the chain of events that lead to metastasis occurrence is urgent to identify new biomarkers of progression and/or new potential targets. Epithelial to mesenchymal transition (EMT) is a major player in cancer dissemination that remains non-targetable due to its broad involvement in tissue homeostasis. Here, we used a relevant *in vitro* dynamic model to identify the pathways triggered during EMT in CRC and how this might improve tumors classification, prognosis and open therapeutic avenue for this deadly disease. The cholesterol pathway surprisingly popped-out from this model and turned out to be a good prognosis factor of disease-free survival for CRC. Altogether, our results showed that an active EMT program lowers the cholesterol pathway, which further influence the differentiation of colon tumor toward the most aggressive subtypes.

## INTRODUCTION

Colorectal cancer (CRC) is the third most common cancer and the second cause of death worldwide (International Agency for Research on Cancer, WHO; http://gco.iarc.fr/today/fact-sheets-cancers). However, the actual therapeutic management proposed to CRC patients is not sufficient. For now, it consists in the surgical removal of the primary tumor, followed by adjuvant chemotherapy in high-risk cases. Metastases are the main cause of death of CRC patients, and thus the major issue in CRC management. Up to 70% of CRC patients will relapse or metastasize within 5 years following surgery of the primary tumor. The 5-year overall survival for metastatic patients is 14% against 90% for non-metastatic patients [1].

Cells prone to metastasis undergo an epithelial-to-mesenchymal transition (EMT), a biological process by which cells are re-programmed, lose their cell-to-cell contacts, polarity and ultimately reshape their cytoskeleton to escape from the initial tumor site [2]. Many works have searched for the initial events that trigger EMT in cancer cells, based on the assumption that targeting EMT inducers might limit metastasis [3, 4]. TGFβ, which is a prominent growth factor at primary tumor sites [5, 6], turned up to be a key inducer of the EMT program [7]. However, TGFβ inhibition was not successful because of the major involvement of this growth factor in many other physiological functions [5, 6]. Moreover, EMT can be triggered in many ways that go beyond the TGFβ pathway alone [8, 9]. We choose to use a combination of different factors known to be involved in the EMT process such as recombinant WNT5A [10, 11], TGFβ [12] and neutralizing antibodies against known inhibitors of EMT, such as SFRP1 [13, 14], DKK1 [15] and CDH1 [16, 17]. The combination of these factors reproducibly induces an EMT phenotype in epithelial cancer cells cultivated *in vitro* within one week: cells detach from their neighbors, adopt spindle shapes and a migratory potential. Characteristic features of treated cells are a decreased expression of adhesion proteins E-Cadherin and Occludin, whereas Fibronectin and Vimentin expression are enhanced [18-20].

The identification of genes specifically activated during the EMT program has been another goal, however difficult to achieve in complex samples such as human tumors. *In vitro* approaches have thus often been privileged. A major common caveat from these approaches was that the cells exposed to the different EMT inducing factorswere usually grown flat [6, 15, 21-23]. We know now that signaling pathways of cells grown in 2D are different from those of tissues and that 3D cultures are more relevant [24, 25]. The extensive study from cells grown in 2D most certainly contributed to limit the identification of EMT regulatory factors to components forming the " tip of the iceberg ", leaving hidden other potential actors. This limitation led us to study how EMT induction can promote pro-invasive features from colon cancer cells, with the aim of revealing novel EMT-related molecular vulnerabilities, potentially useful as new prognostic factors and/or novel targets.

For this, we used a 3D *in vitro* model and a timeline exposition to the EMT-inducing cocktail to study reversible genes involved in EMT and Mesenchymal-to-Epithelial Transition (MET) programs. From the genes identified, we build a metagene and tested its prognostic value in a large database of CRC samples, annotated with clinical data. Finally, still using these genes, we searched for FDA approved drugs that might influence the EMT-MET programs and found that statins alter the execution of the EMT program.

## MATERIAL& METHODS

### Cell culture

HT29 cells were obtained from the ATCC^®^ Human Cell Lines (ATCC, Manassas, VA, USA) and cultured using Dulbecco Modified Eagle Medium (DMEM, GIBCO, Waltham, MA, USA) with 10% Fetal Calf Serum (FCS, Eurobio, Les Ulis, France) in 5% CO_2_ at 37°C. Classical 2D cell cultures were seeded in Falcon^®^ 6-well, whereas spheroids culture were seeded in Cell Carrier Spheroid ULA 96-well Microplates™ (PerkinElmer, Waltham, MA, USA). For EMT induction, cells were platted 2 days in regular media before to be supplemented with StemXVivo^®^ EMT Inducing Media Supplement (R&D, Minneapolis, MN, USA) at 1X concentration for the indicated time. Cells were treated with lovastatin and simvastatin (Sellekchem, Houston, TX, USA) at indicated concentration and time.

### Western Blotting

Cells were scraped and lysed in RIPA buffer (150 mM NaCl, 50 mM Tris, pH 7.4, 1% Triton X-100, 0.1% SDS, 1% sodium deoxycholate) supplemented with HALT protease inhibitor 1X (Thermo Fisher Scientific, Waltham, MA, USA), incubated for 10 min on ice, and centrifuged at 13800 g. Supernatants were collected and quantified with Bradford Protein Assay kit (Thermo Fisher Scientific, Waltham, MA, USA). NuPAGE^®^ LDS Sample Buffer (Invitrogen, Carlsbad, CA, USA) was added to samples to 1X final concentration. Samples were boiled at 95 °C for 5 min, loaded into acrylamide/bisacrylamide gels and transferred to nitrocellulose membrane (GE Healthcare, Chicago, IL, USA). Membranes were blocked using PBS 0.1% tween 5% fat free milk for at least 30 min and incubated with primary antibodies diluted in PSB 5% normal horse serum overnight at 4 °C (**Table S1**). The following day, membranes were incubated with HRP coupled secondary antibodies diluted in PSB 5% normal horse serum for 1 h at room temperature (**Table S1**). Blots were visualized by addition of ECL Western Blotting Substrate (Thermo Fisher Scientific, Waltham, MA, USA) following manufacturer’s instructions. Blots were revealed using the imaging system ChemiDoc MP (BioRad, Hercules, CA, USA).

### RNA extraction

Total RNA was extracted from the HT29 spheroids using the RNeasy Mini Kit (Qiagen, Hilden, Germany) following the manufacturer’s instruction. The quality was tested using an Agilent Bioanalyzer (Agilent, Santa Clara, CA, USA).

### Gene expression profiling of treated and untreated HT29 spheroids

Profiling was done using Affymetrix GeneChip HuGene 2.0 ST microarrays according to manufacturer instructions (Affymetrix, Thermo Fisher, Waltham, MA, USA). Expression data were analyzed by Robust Multichip Average (RMA) [26] in R using Bioconductor and associated packages. Prior to analysis, log2-transformed data were filtered out to exclude poorly expressed genes, defined as genes having a median expression level across all samples lower than the first quartile range. Data filtering resulted in 17,421 annotated genes.

Unsupervised analysis, applied to the 3,131 genes with standard deviation (SD)>0.25, was performed with the Cluster program [27] using Pearson correlation and average linkage clustering as parameters. Results were displayed using TreeView program [27]. Robustness of clusters was assessed with the R-package pvclust [28] with identical clustering parameters and 100 bootstrap replications. Ontology analysis of the gene clusters was based on GO biological processes of the Database for Annotation, Visualization and Integrated Discovery (DAVID; http://david.abcc.ncifcrf.gov/). The Consensus Molecular Subtype (CMS) classification [29] was applied to each cell culture condition by using the CMScaller tool from Eide et al. [30], providing a CMS score for each subtype (CMS1, CMS2, CMS3, and CMS4) to each condition.

Supervised analyses between the different cell culture conditions was applied by using Gene Set Enrichment Analysis (GSEA) (http://www.broadinstitute.org/gsea/) in order to identify the differential biological pathways. Analysis was focused on the 50-hallmark gene sets from the MSigDB v7.0 Molecular Signatures Database (https://www.gsea-msigdb.org/gsea/msigdb). We used the class differential metric to rank the filtered genes, weighted enrichment statistic for computing enrichment score (ES) of each gene set tested and 1000 permutations to evaluate significance as parameters for the GSEA. Significant gene sets were defined by a False Discovery Rate (FDR)-corrected p-value <0.01. Metagenes from the significant gene sets were computed as the mean expression of core genes defined by the GSEA analysis.

### Gene expression profiling of human cancer colon samples

We built a large database gathering our own data set and 10 publicly available sets as described [26]. These latter had been collected from the National Center for Biotechnology Information (NCBI)/Genbank GEO, ArrayExpress and TCGA databases (**Table S2**). Samples had been profiled using DNA microarrays (Affymetrix, Thermo Fisher, Waltham, MA, USA) or Illumina RNA sequencing (United kingdom, Cambridge). The whole data set included 2,239 primary colon cancer samples in the final analysis. Their clinicopathological characteristics are summarized in **Table 1**. The pre-analytic data processing and data analysis were done as previously described [26]. The CMS classification and the metagenes identified by supervised analyses in the HT29 cells were applied to each data set separately [29]. For these latter, the natural score of 0 was used as threshold to define a sample as “positive” (score>0) or “negative” (score<0).

**Table 1:** Clinicopathological characteristics of the 2,239 primary colon cancer samples

| Characteristics |  | N (%) |
| --- | --- | --- |
| Age_Diag50 | <=50 | 228 (12%) |
|  | >50 | 1702 (88%) |
| Sex | female | 954 (47%) |
|  | male | 1069 (53%) |
| Location | distal | 840 (50%) |
|  | proximal | 854 (50%) |
| pStage | 1 | 167 (10%) |
|  | 2 | 667 (41%) |
|  | 3 | 548 (34%) |
|  | 4 | 237 (15%) |
| Grade | 1 | 26 (6%) |
|  | 2 | 353 (77%) |
|  | 3 | 77 (17%) |
| CMS | CMS1 | 389 (20%) |
|  | CMS2 | 640 (32%) |
|  | CMS3 | 343 (17%) |
|  | CMS4 | 604 (31%) |
| Follow-up median, months (range) |  | 43 (1-212) |
| 5y-RFS |  | 71% [68-73] |

### Connectivity Map interrogation

Finally, in order to identify potential therapies, we applied the Connectivity Map (CMap) analysis [31]. This analysis contains a perturbation-driven gene expression dataset of over a million of gene expression signatures associated with molecule compounds and genetic perturbations treatment on human cells. CMap (https://clue.io/cmap) searched for, among the L1000 signatures, the perturbagen’s signatures that give significant correlation with those genes included in our two interest metagenes given by connectivity scores. The tested signatures were limited to those with a well-defined mechanism of action of treatment compounds, corresponding to 10,847 signatures of 3,248 drugs through 57 cultured human cell lines. The significance threshold of the normalized connectivity score (ncs) was adjusted p-value <0.001. Because each drug has been used through multiple different culture conditions, we aggregated the significant signatures by drug class, computed for each class the average of normalized connectivity scores (average ncs) and assessed its statistical significance using one sample Student t-test.

### Statistical analysis

Correlations between the “EMT-CHL model”-based classes and the clinicopathological variables were calculated with the Student’s t-test for the continuous variables and the Fisher’s exact test for the binary variables. The survival endpoint was relapse-free survival (RFS) calculated from the date of diagnosis until the date of relapse. The follow-up was measured from the date of diagnosis to the date of last news for event-free patients. Survival was calculated using the Kaplan-Meier method and curves were compared with the log-rank test. Univariate and multivariate analyses were done using Cox regression analysis (Wald test). The variables tested in univariate analysis included patients’ age (> *vs*. ≤50 years) and gender (male *vs*. female), anatomic location (proximal *vs*. distal colon), pathological stage (2, 3, 4 *vs*. 1), pathological grade (2, 3 *vs*. 1), CMS molecular subtypes (CMS2, CMS3, CMS4 *vs*. CMS1), and the “EMT-CHL model”-based classes (“high-risk” *vs*. “low-risk”). Multivariate analysis incorporated all variables with a p-value inferior to 5% in univariate analysis. The likelihood ratio (LR) tests were used to assess the prognostic information provided beyond that of each metagene included in the Cox model, assuming a X2 distribution. Changes in the LR values (LR-ΔX2) quantified the relative amount of information of one Cox model compared with another. In order to build a prognostic Cox model for RFS combining our two metagenes, we randomly split the 1,837 informative primary samples in two sets, training and validation sets. The training set was composed of a quarter of the samples (n=450) and allowed to build the model. This model was then applied to the validation set composed of the 1,387 remaining samples, to estimate the robustness of the predictor. All statistical tests were two-sided at the 5% level of significance. Statistical analysis was done using the survival package (version 2.30) in the R software (version 3.5.2).

## RESULTS

### The EMT-inducing cocktail induces reversible loose micro-tissues, consistent with EMT and EMT-MET-like programs

The EMT-inducing cocktail used for this study is a potent inducer of EMT [18, 20, 32-35]. However, growth in 2D on plastic dishes offers unnatural growth kinetics and cells attachment, thus altering adhesion and related signaling pathways, and potentially biasing and/or masking EMT-induced signals. To study EMT induction in a more physiologic and dynamic model, we looked at the reversible effects of an EMT-inducing cocktail on cells grown in 3D as spheroids. We induced EMT on several colon cancer cell lines (HT29, HCT116, SW480). The most striking phenotype was observed with the HT29 colon cancer cell line, and we choose to continue with them. We used three conditions on HT29 spheroids: HT29 spheroids grown without any treatment (Baseline condition), HT29 spheroids exposed to the EMT-inducing cocktail for 5 days to induce EMT, and HT29 spheroids exposed to the EMT-inducing cocktail for 5 days, washed, then incubated for an additional 4 days without any treatment to mimic a sequential EMT followed by a MET process (**Figure 1A**).

**Figure 1:**
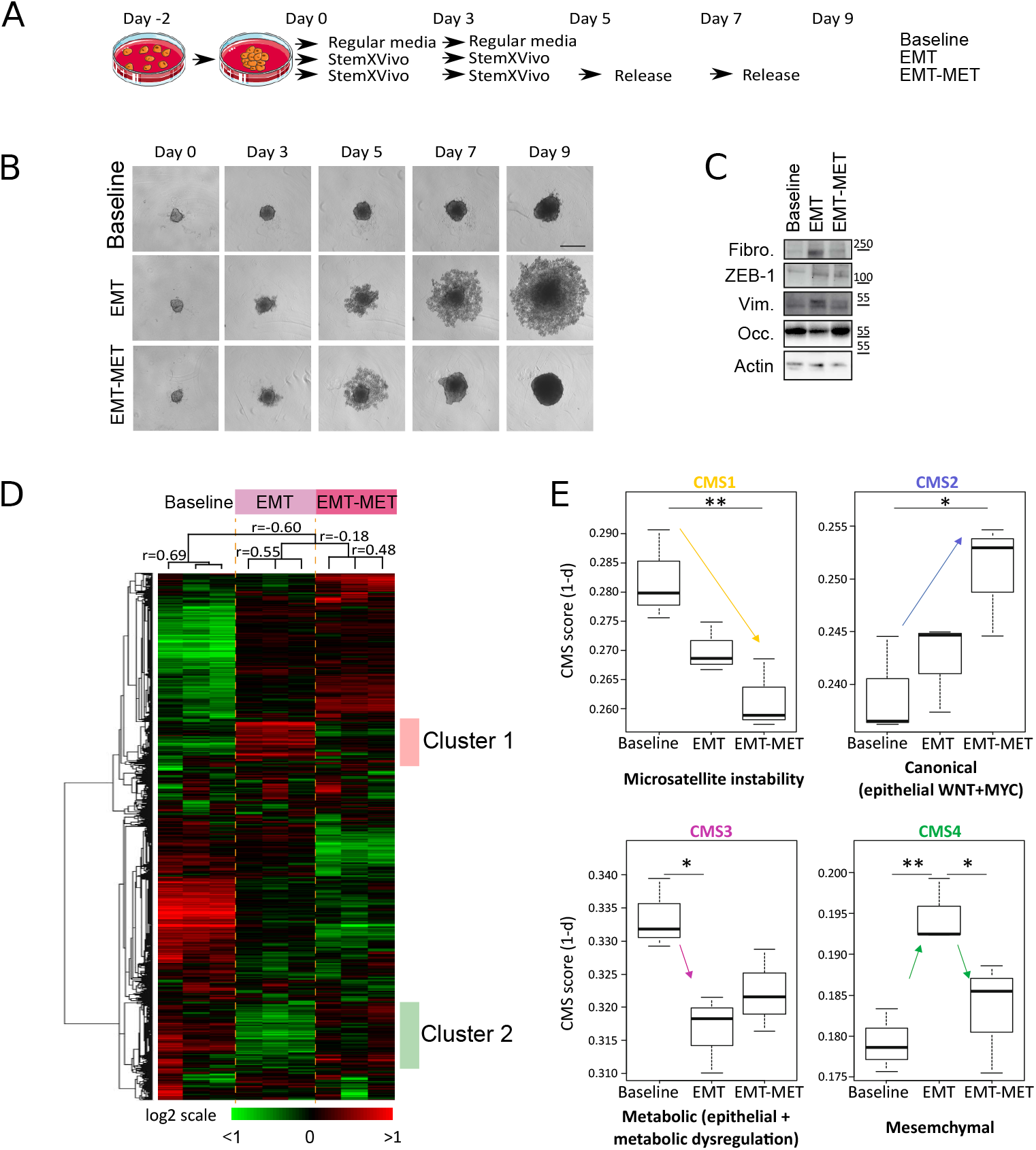
Characterization and validation of the EMT induced model. Cells were grown in 3D in ULA plate for 2 days (Day-2). At day 0, the EMT and EMT-MET samples were treated with the EMT-inducing cocktail for 5 days. For EMT-MET samples, the spheroids were washed off the EMT-inducing cocktail and grown for 4 more days in normal medium. **A)** Experimental setting. **B)** Representative photos of the spheroids for the baseline, EMT, EMT-MET conditions, at days 0, 3, 5, 7 and 9. **C)** Western Blots of proteins involved in EMT: FIBRONECTIN, ZEB1, VIMENTIN, OCCLUDIN. ACTIN was used as loading control. **D)** Hierarchical clustering of triplicate samples of three conditions i.e. baseline, EMT and EMT-MET and 3,131 genes based on mRNA expression levels with standard deviation (SD)>0.25. Each row represents a gene and each column represents a sample. Robust clusters defined by pvclust with an approximately unbiased p-value>100% and identified with (an asterisk and) the associated Pearson correlation. **E)** The Consensus Molecular Subtype (CMS) classification was applied to mRNA expression profile of each sample by using the CMScaller tool from Eide et al. [21], providing a score for the four CMS subtypes (CMS1, CMS2, CMS3, and CMS4). Box plot comparing the four CMS scores according each condition (baseline, EMT or EMT-MET). The significant differences defined by one-way ANOVA post hoc Tukey test are visualized with an arrow and p-value are indicated as follows. * : p < 0.05, **: p < 0.01

In the absence of any specific treatment (Baseline), the spheroids continued to grow as dense spheroids during the whole protocol (**Figure 1B, top row**). When the EMT-inducing cocktail was added to the media (EMT condition) the spheroids became loose, with round cells at the periphery of the spheroid core, already after 3 days of treatment (**Figure 1B, middle row**). In the EMT-MET condition, the first 5 days were similar to the EMT condition, then during the following 3 days, *i*.*e*. after the removal of the EMT-inducing cocktail (MET-*like* program), the individualized round cells progressively reintegrated the spheroids core to form a homogeneous and compact spheroid (**Figure 1B, bottom row**).

We verified that the EMT-like phenotype induced by the EMT-inducing cocktail actually induced known EMT factors. We studied key molecules of EMT at the protein level by western blotting. We compared Baseline microtissues samples to EMT and EMT-MET samples. The EMT-inducing cocktail induced the expression of specific mesenchymal proteins such as Fibronectin, ZEB1 and Vimentin (**Figure 1C)**. All these markers went back to their respective baseline after removal of the EMT-inducing cocktail (*i*.*e*. Baseline condition and EMT-MET are similar). Conversely and as expected, the epithelial marker OCCLUDIN was downregulated during the EMT process.

Thus the EMT-inducing cocktail induced reversible macroscopic changes, phenotypically compatible with an EMT program induction.

### Transcriptomic analysis confirms the activation of EMT and EMT-MET programs in spheroids treated with the EMT-inducing cocktail

We surmised that the effect of the EMT-inducing cocktail was not limited to known EMT markers. To identify broad biological pathways activated by the EMT-inducing cocktailin treated spheroids, we performed DNA microarray analyses (Affymetrix™ GeneChip Human Gene 2.0 ST Assay) from the three conditions described above (Baseline, EMT, and EMT-MET, **Figure 1A**) and compared their expression profiles. Analyses were done at day 5 for Baseline and EMT samples and at day 7 for EMT-MET samples. Each condition was tested in triplicate. Unsupervised hierarchical clustering revealed, as expected, three distinct and coherent clusters, corresponding to each of the three conditions tested: Baseline, EMT, and EMT-MET (**Figure 1D**). The EMT and EMT-MET conditions were closer to each other than to the Baseline condition (**Figure 1D**). This is coherent with the fact that at day 7, spheroids were not fully back to baseline yet (**Figure 1B**). Visual inspection of gene clusters revealed two clusters, thereafter designated 1 and 2, that were deregulated from the Baseline condition to the EMT condition and showed opposite deregulation from the EMT condition to the EMT-MET condition. Cluster 1 was upregulated in EMT condition and downregulated in EMT-MET condition: it included 198 genes with minimal Pearson correlation equal to 0.74. In term of ontologies, the DAVID GO biological processes associated with this cluster included cell migration (p=1.15E-04), extracellular matrix organization (p=1.78E-04), regulation of cell adhesion (p=7.87E-05) and wound healing pathways (p=3.84E-03). By contrast, Cluster 2 was downregulated in EMT condition and upregulated in EMT-MET condition: it included 296 genes with minimal Pearson correlation equal to 0.75. The GO biological processes represented in this cluster were related to cholesterol biosynthetic process (p=4.42E-20), lipid metabolism (1.6E-02) and metabolic pathways (p=1.22E-02) (**Figure 1D, Table S3)**. Thus, transcriptional analysis of the EMT and EMT-MET programs induced in the spheroids revealed a list of candidates and associated molecular pathways whose expression was modulated and reversible. Expected pathways related to cell migration and adhesion, known to be central to this reversible phenomenon, validated the biological relevance of our *in vitro* model that also revealed unexpected genes and pathways.

### Spheroids treated with the EMT-inducing cocktail shift toward the mesenchymal consensus molecular subtypes (CMS4)

The CMS classification distinguishes four subtypes of CRC based on the tumor’s prominent biological profiles, each subtype having a defined prognosis: the CMS1 microsatellite instability/immune subtype (hyper-mutated, microsatellite unstable and strong immune activation), the CMS2 canonical subtype (epithelial with marked WNT and MYC signaling activation), the CMS3 metabolic subtype (epithelial and neat metabolic dysregulation), and the CMS4 mesenchymal subtype (prominent TGFβ activation, stromal invasion and angiogenesis) [36]. We surmised that genes expressed in the CMS4 mesenchymal class may be enriched in the EMT-inducing cocktail treated spheroids, but neither in the Baseline nor in the EMT-MET spheroids.

The CMS classification applied to the 3 cell culture conditions (Baseline, EMT and EMT-MET conditions) defined a CMS score for each subtype (CMS1, CMS2, CMS3, and CMS4) and for each cell culture condition (Baseline, EMT and EMT-MET conditions). As shown in **Figure 1E**, the prevalent score of Baseline samples corresponded to the CMS3 class (mean CMS3 score = 0.332). This result is coherent with a previous CMS classification of HT29 cells [37]. When compared with Baseline conditions, EMT samples showed a decrease of the CMS3 score (p=1.22E-02) and an increase of the CMS4 score (p=9.58E-03). Enrichment in CMS4 score (*i*.*e*. mesenchymal genes) was concordant with the induction of EMT. After the EMT-inducing cocktail removal (EMT-MET samples), the score for the CMS4 class decreased, as expected, while the score for the CMS2 class increased compared to the Baseline condition (p=2.57E-02). This result showed that a pro-epithelial program (starting with a switch toward the CMS2 subtype) was re-activated once the EMT-inducing cocktail was removed from the culture. Altogether, the results showed that HT29 cells grown as spheroids and exposed to an EMT inducer activated a physiologic pro-mesenchymal transcriptomic program (the shift toward the CMS4 mesenchymal subtype) and that this process was reversible toward a pro-epithelial phenotype when the EMT inducer was removed. This observation further validated the relevance of our *in vitro* model.

### EMT genes, Bile Acid Metabolism and Cholesterol Homeostasis hallmark gene sets are affected during the EMT and EMT-MET programs

To identify the biological pathways reversibly altered during the EMT process, we did a supervised analysis between the different cell culture conditions. Two comparisons were done: EMT *versus* Baseline samples (EMT induction), and EMT-MET *versus* EMT samples (MET induction). From these signatures, we used GSEA applied to the 50-hallmark gene sets from the MSigDB Molecular Signatures Database. Our objective was to identify and select gene sets that showed an opposite pattern of variation in those two comparisons. Sixteen gene sets were differentially regulated at least in one comparison (q-value<1%) (**Figure 2A**, *grey background*), and three of them were significantly up-or downregulated in the EMT samples and returned to baseline levels in the EMT-MET samples (**Figure 2A**, *white background*). The “*Epithelio-Mesenchymal transition*” gene set was composed of 23 core genes upregulated in the EMT compared to Baseline samples (q-value = 5.61E-03) and 39 core genes downregulated in the EMT-MET compared to EMT samples (q-value < 1.00E-03) (**Table S4**, *columns 3 and 10 respectively, orange background*). The *Bile Acid Metabolism* gene set was composed of 33 core genes downregulated in the EMT compared to Baseline samples (q-value = 9.16E-03) and 33 core genes upregulated in the EMT-MET compared to EMT samples (q-value = 1.62E-03) (**Table S4**, *columns 3 and 10 respectively, green background*). The *Cholesterol Homeostasis* gene set was composed of 31 core genes downregulated in the EMT samples compared to Baseline samples (q-value <1.00E-03) and 28 core genes upregulated in the EMT-MET compared to EMT samples (q-value= 2.64E-03) (**Table S4**, *columns 3 and 10 respectively, yellow background*). Among the core genes: 14 genes were retained in the “*Epithelio-Mesenchymal transition”* gene set, 17 in the “*Bile Acid Metabolism”* gene set, and 21 in the “*Cholesterol Homeostasis”* gene set, representing a total of 52 genes (**Figure 2B, Table S4** *column 18*).

**Figure 2.**
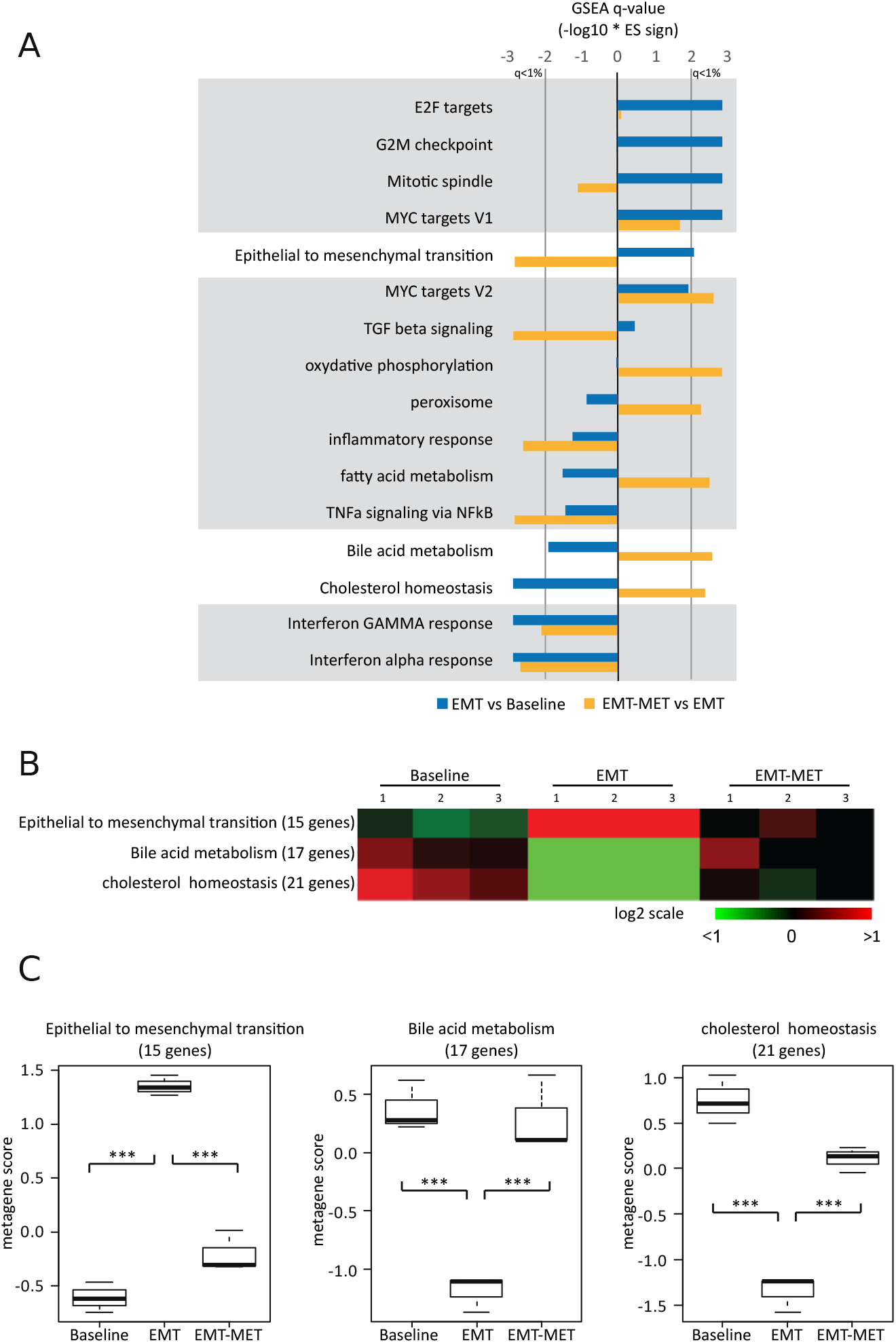
Epithelial to Mesenchymal transition pathways, Cholesterol homeostasis and bile acid metabolism are inversely regulated by the addition and the release of the EMT-inducing cocktail. **A)** Sixteen gene-sets out from the MSigDB Molecular Signatures Database (GSEA) were differentially regulated in at least one comparison (q-value<1%): Baseline vs EMT or EMT vs EMT-MET. The 3 gene sets inversely regulated in the two conditions were “*Epithelial Mesenchymal Transition”*; “*Bile Acid Metabolism*”; “*Cholesterol Homeostasis”* (white background). **B**) Heatmap of metagenes in the nine samples ordered by condition i.e. Baseline, MT and EMT-MET conditions. Metagene of the three gene sets that were significantly and oppositely deregulated in both comparisons were built using the core genes defined in GSEA analysis: “*Epithelial Mesenchymal Transition”* (14 genes); “*Bile Acid Metabolism”* (17 genes); “*Cholesterol Homeostasis”* (21 genes). **C)** Box plot comparing the three individual metagenes according each condition (baseline, EMT or EMT-MET). P-values (ANOVA post hoc Tukey test) are indicated as follows *** p < 0.001.

We built 3 independent metagenes from each of these three core gene sets. As shown in **Figure 2C**, each metagene was robust enough to distinguish the HT29 samples according to their respective condition (Baseline, EMT, and EMT-MET): the “*Epithelio-Mesenchymal transition”* metagene score was the highest in the EMT samples compared to the Baseline samples (p-value=3.22E-06) and to the EMT-MET samples (p-value=1.25E-05). The “*Bile Acid Metabolism”* and the “*Cholesterol Homeostasis”* metagene scores were the lowest in the EMT samples compared to the Baseline samples (p-value=2.08E-04 and p-value=1.70E-05 respectively) and to the EMT-MET samples (p-value=2.08E-04 and p-value=1.70E-05, respectively). Altogether, this procedure allowed us to identify 52 genes reversibly regulated during the EMT and the EMT-MET programs. Those genes, when separated into three distinct metagenes based on their hallmark ontology, could also individually distinguished an active from an inactive EMT program in tumor cells.

### *In vitro* derived metagenes show independent prognostic value in colon cancer

Given the known link between EMT and the metastatic process, we assessed the prognostic value of each metagene (positive *versus* negative) in a large gene expression database of untreated human colon cancer samples (**Table S2**). This base included 2,239 clinically annotated primary colon tumors, including 1,837 with available Recurrence-Free Survival (RFS) data. In univariate analysis, the “*Bile Acid Metabolism”* metagene was not prognostic (**Figure 3A**). By contrast, the positive “*Epithelio-Mesenchymal transition”* metagene (HR=1.37, [95CI 1.14-1.64]; p=6.28E-04, Wald test) and negative “*Cholesterol Homeostasis”* metagene (HR=1.25, [95CI 1.04-1.49]; p=1.64E-02, Wald test) were associated with shorter RFS, and showed independent prognostic value in multivariate analysis (**Figure 3B**). The **Figure 3C** shows the Kaplan-Meier curves for RFS in four groups defined by each significant metagene: the 5-year RFS was 77% (95%CI 73-82) in the “*Epithelio-Mesenchymal transition”-* negative/”*Cholesterol Homeostasis”-*positive group, 71% (95%CI 67-77) “*Epithelio-Mesenchymal transition”-*negative/”*Cholesterol Homeostasis”-*negative group, 70% (95%CI 65-75) in the “*Epithelio-Mesenchymal transition”-*positive/”*Cholesterol Homeostasis”-*positive group, and 65% (95%CI 60-70) in the “*Epithelio-Mesenchymal transition”-*positive/”*Cholesterol Homeostasis”-*negative group (p=4.61E-04; log-rank test). Such prognostic complementarity was confirmed using the likelihood ratio (LR) test (**Table S5**): the “*Cholesterol Homeostasis”* metagene added prognostic information to that provided by “*Epithelio-Mesenchymal transition”* metagene (ΔLR-X2=5.21, p=2.24E-02), and the “*Epithelio-Mesenchymal transition”* metagene added prognostic information to that provided by the “*Cholesterol Homeostasis”* metagene (ΔLR-X2=13.26, p=2.72E-04).

**Figure 3:**
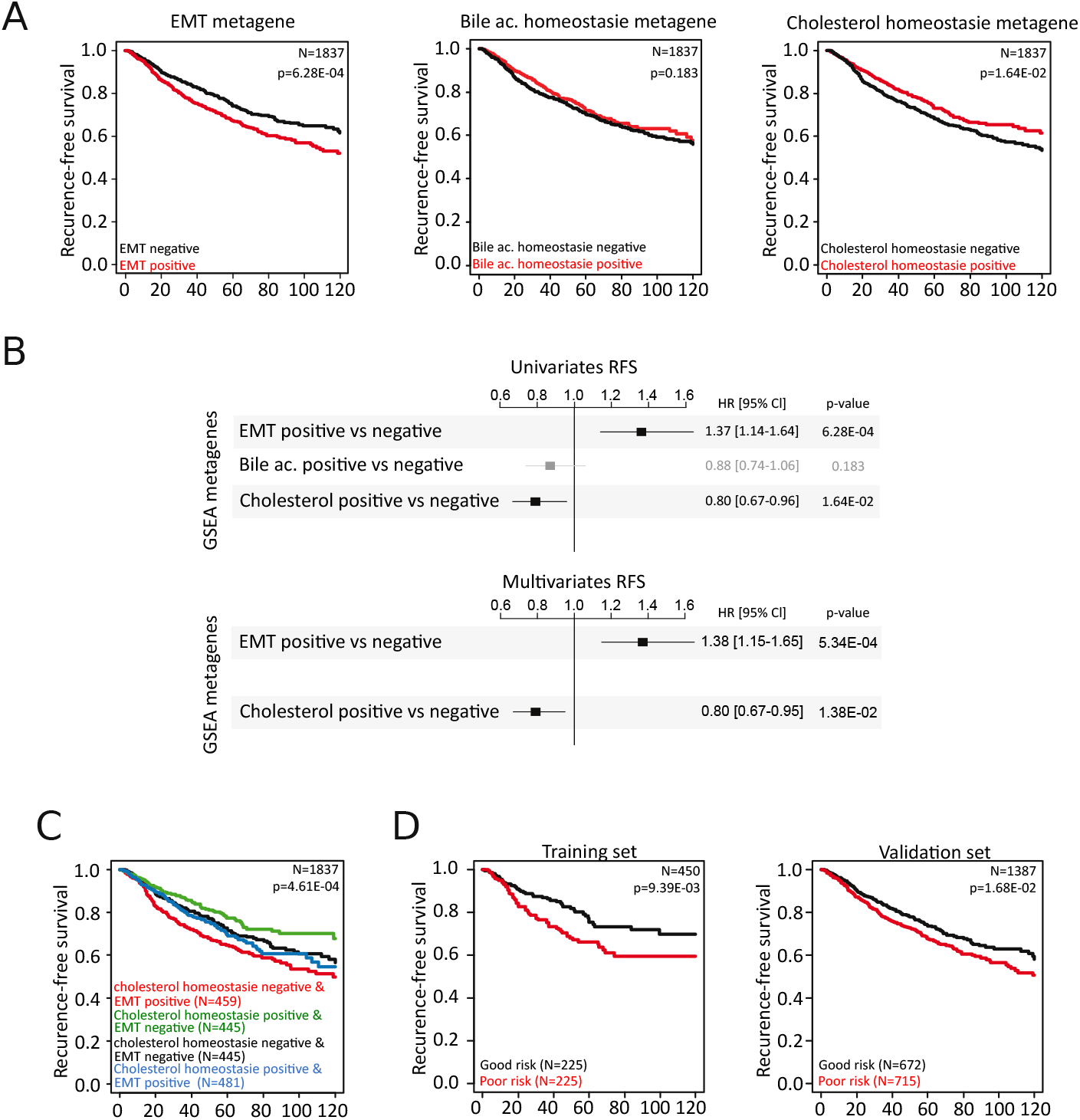
Epithelial to Mesenchymal transition pathways and Cholesterol homeostasis have synergic prognostic value in patients. Patients were classified as positive or negative according to their metagenes scores. **A)** Kaplan-Meier curves for recurrence free survival (RFS) of 1887 CRC patients assessed using the log-rank test, for the 3 metagenes: “*Epithelial Mesenchymal Transition”*, “*Bile Acid Metabolism”* and “*Cholesterol Homeostasis”*. **B)** Forest plot showing the hazard ratio for survival events of each GSEA metagene and recurrence-free survival in CRC patients in univariate and mutivariante analysis. A ratio greater than one indicates a poor prognosis and a ratio lower than one indicates good prognosis. The black squares correspond to significant genes and the grey ones to non-significant and the lines defined the 95% confidence interval. **C)** Kaplan-Meier curves for RFS according to four groups: *Epithelio-Mesenchymal transition”-*negative/”*Cholesterol Homeostasis”-*positive” group (green, n=452), “*Epithelio-Mesenchymal transition”-*negative/”*Cholesterol Homeostasis”-*negative group (black, n=445), “*Epithelio-Mesenchymal transition”-*positive/”*Cholesterol Homeostasis”-*positive group (blue, n=481), and “*Epithelio-Mesenchymal transition”-*positive/”*Cholesterol Homeostasis”-*negative group (red, n=459). ***D)*** Kaplan-Meier curves for survival model derived from the fusion of and Cholesterol Homeostasis metagenes (“*Epithelial Mesenchymal Transition”* and “*Cholesterol Homeostasis”*.) in the training (n=450) and the validation (n=1387) sets.

#### The model combining the “Epithelial Mesenchymal Transition” and the “Cholesterol Homeostasis” metagenes is a robust prognostic factor for RFS in CRC

Based on these results, we built a multigene model combining the “*Epithelial Mesenchymal Transition*” (14 genes) and the “*Cholesterol Homeostasis*” (21 genes) metagenes, thereafter designated “EMT-CHL model”. We randomly split the 1,837 informative primary samples in two sets: a training set composed of a quarter of the samples (n=450) to build the model, and a validation set composed of the 1,387 remaining samples (**Table 2**) to test its robustness. As expected, in the training set, the model showed a difference between the 5-year RFS of the “low-risk” class (75% [CI95 69-83]; n=225) and the “high-risk” class (66% [CI95 59-74]; n=225; p=9.93E-03, log-rank test; **(Figure 3D**, *left panel*). Importantly, the model maintained its prognostic value in the validation set, with 74% 5-year RFS in the “low-risk” class (CI95 70-78; n=672) *versus* 68% in the “high-risk” class (CI95 64-72; n=71; p=1.68E-23, log-rank test) **(Figure 3D**, *right panel*).

**Table 2:** Correlations of the “EMT-CHL model”-based classes with clinicopathological variables

| EMT' + 'CHL hom.' Cox model<br>(Validation set) |  |  |  |  |
| --- | --- | --- | --- | --- |
| Characteristic | N | low-risk | high-risk | p-value |
| Age_Diag50 |  |  |  | 0.0507 |
| <=50 | 139 | 25 (8%) | 114 (13%) |  |
| >50 | 1064 | 277 (92%) | 787 (87%) |  |
| Sex |  |  |  | 0.00201 |
| female | 603 | 127 (40%) | 476 (50%) |  |
| male | 668 | 192 (60%) | 476 (50%) |  |
| Location_2K |  |  |  | 0.921 |
| distal | 517 | 130 (48%) | 387 (49%) |  |
| proximal | 543 | 139 (52%) | 404 (51%) |  |
| pStage |  |  |  | 3.00E-04 |
| 1 | 123 | 34 (15%) | 89 (12%) |  |
| 2 | 466 | 130 (56%) | 336 (44%) |  |
| 3 | 399 | 67 (29%) | 332 (44%) |  |
| 4 | 0 | 0 (0%) | 0 (0%) |  |
| Grade |  |  |  | 0.876 |
| 1 | 19 | 5 (7%) | 14 (6%) |  |
| 2 | 259 | 59 (78%) | 200 (80%) |  |
| 3 | 47 | 12 (16%) | 35 (14%) |  |
| CMS |  |  |  | 3.50E-33 |
| CMS1 | 269 | 52 (19%) | 217 (23%) |  |
| CMS2 | 401 | 138 (51%) | 263 (28%) |  |
| CMS3 | 212 | 79 (29%) | 133 (14%) |  |
| CMS4 | 343 | 3 (1%) | 340 (36%) |  |
| Follow-up median, months (range) | 1387 | 42 (1-192) | 42 (1-203) | 0.387 |
| 5y-RFS | 1387 | 74% [70-78] | 68% [64-72] | 1.68E-02 |

The correlation of the “EMT-CHL model”-based classes with clinicopathological variables in the validation set are shown in **Table 2**. When compared with the “low-risk” class, the “high-risk” class was associated with more female (p=2.0E-03, Fisher’s exact test), more pathological stage 3 (p=3.0E-04, Fisher’s exact test), and more CMS1 and CMS4 subtypes (p=3.5E-33, Fisher’s exact test). We then compared the prognostic value of our “EMT-CHL model” with that of other clinicopathological factors in the validation set in univariate and multivariate analyses (**Table 3**). The other variables significant in univariate analysis were the pathological stage (p=3.1E-10) and the CMS classification (p=2. 3E-03, Wald test). In multivariate analysis, the pathological stage and our multigene model (p=4.0E-02, Wald test) remained significant but not the CMS classification. Altogether these analyses showed that our “EMT-CHL” model was a robust and independent prognostic factor for RFS in primary colon cancer.

**Table 3:**
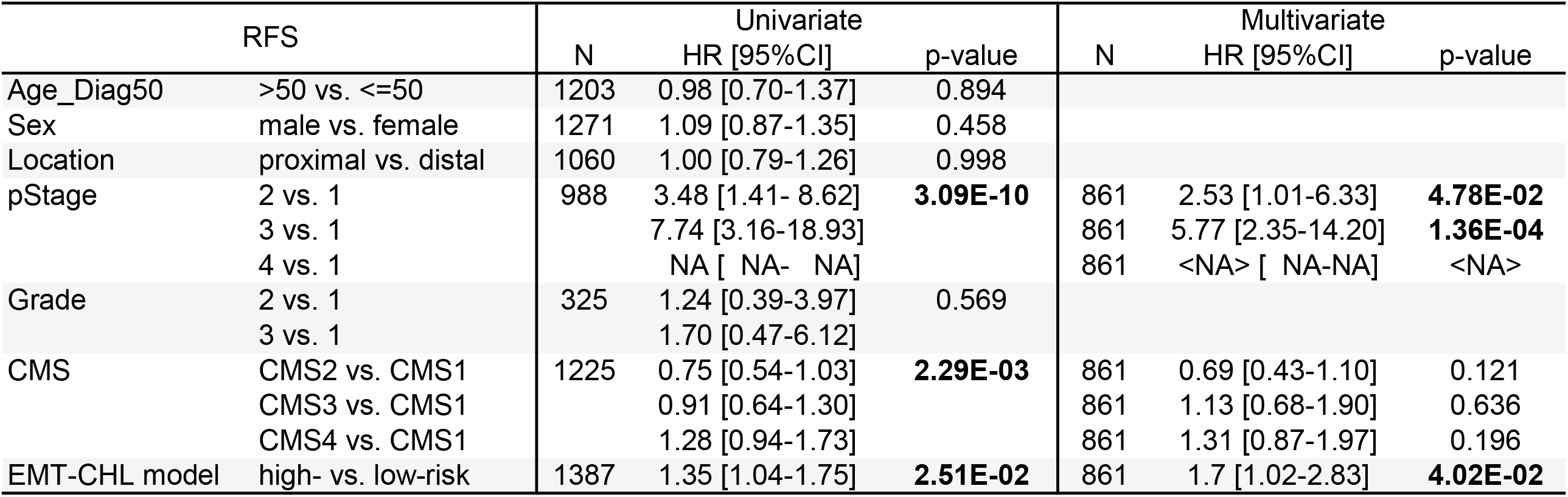
Uni- and multivariate analyses for RFS in the validation set

#### CMap analysis revealed inhibitors and antagonists that might affect the EMT and EMT-MET programs

We then searched for drugs that might affect (promote or inhibit) the plasticity of the EMT and/or EMT-MET programs. We submitted the 35 genes of the EMT-CHL model to Connectivity Map (CMap), which reports the transcriptional responses of human cells to a variety of chemical or genetic perturbations [31]. The resulting connectivity scores reflect the level of agreement between the tested signature and the L1000 profiles/signatures. Analysis was limited to the 10,847 signatures related to treatment compounds with known mechanism of action through 57 cultured human cell lines: 761 signatures had significant correlation (normalized connectivity score (ncs) *i*.*e*. adjusted p-value <0.001) with the “EMT-CHL” model genes through 47 human cell lines over 309 drugs compounds (representing 145 drug classes; **Table S6**). We aggregated the significant signatures by drug class and computed the average ncs for each class: 98 drug classes were significant. **Table 4** shows the top 15 drug classes identified, which includes antagonists/inhibitors of Dopamine receptor, EGFR, Serotonin receptor, PDGFR, BCR-ABL, ABL, Histamine receptor, KIT, Adrenergic receptor, HMG-CoA reductase (HMGCR), SRC, Calcium channel, Tricyclic antidepressant, Estrogen receptor, and VEGFR. All connectivity scores were negatively correlated with our signature, suggesting that these drugs might interfere with the EMT-MET process.

**Table 4:**
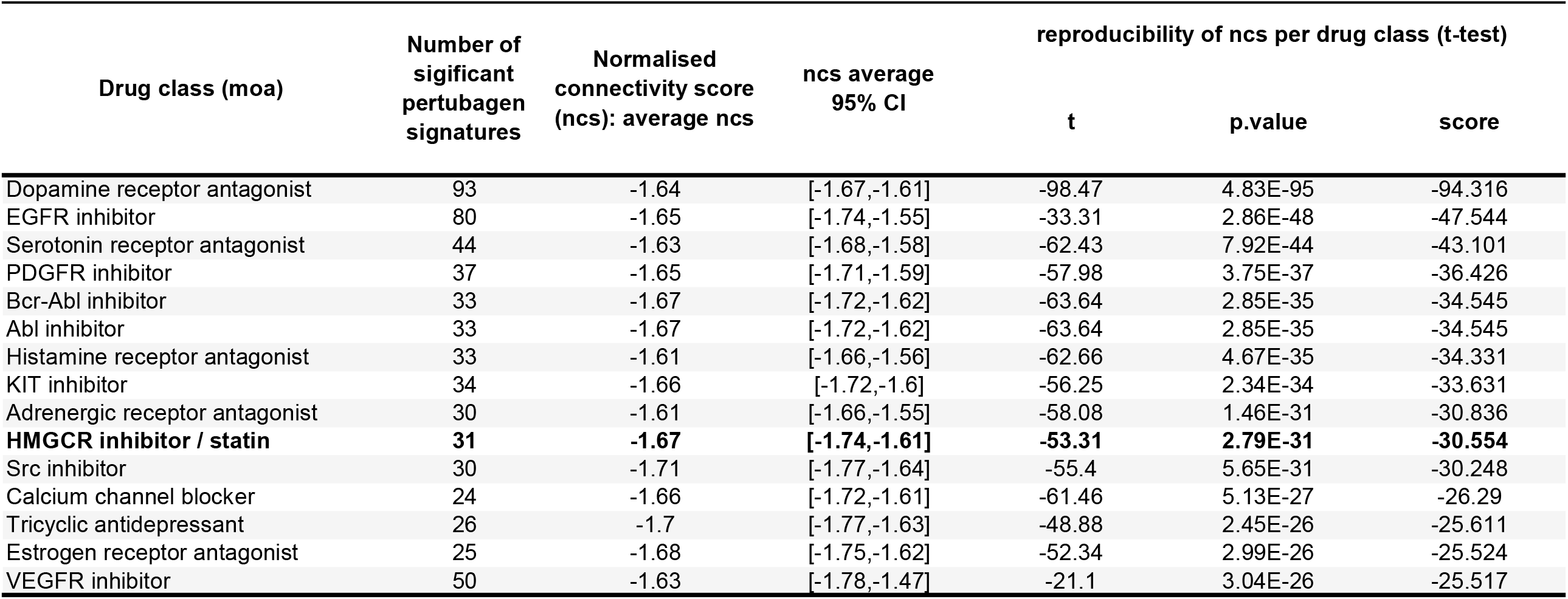
Top 15 drug classes identified with connectivity map (L1000), 14 EMT vs. 21g CHL GSEA core genes

| Drug class (moa) | Number of significant pertubagen signatures | Normalised connectivity score (ncs): average ncs | ncs average 95% CI | reproducibility of ncs per drug class (t-test) |  |  |
| --- | --- | --- | --- | --- | --- | --- |
|  |  |  |  | t | p.value | score |
| Dopamine receptor antagonist | 93 | -1.64 | [-1.67,-1.61] | -98.47 | 4.83E-95 | -94.316 |
| EGFR inhibitor | 80 | -1.65 | [-1.74,-1.55] | -33.31 | 2.86E-48 | -47.544 |
| Serotonin receptor antagonist | 44 | -1.63 | [-1.68,-1.58] | -62.43 | 7.92E-44 | -43.101 |
| PDGFR inhibitor | 37 | -1.65 | [-1.71,-1.59] | -57.98 | 3.75E-37 | -36.426 |
| Bcr-Abl inhibitor | 33 | -1.67 | [-1.72,-1.62] | -63.64 | 2.85E-35 | -34.545 |
| Abl inhibitor | 33 | -1.67 | [-1.72,-1.62] | -63.64 | 2.85E-35 | -34.545 |
| Histamine receptor antagonist | 33 | -1.61 | [-1.66,-1.56] | -62.66 | 4.67E-35 | -34.331 |
| KIT inhibitor | 34 | -1.66 | [-1.72,-1.6] | -56.25 | 2.34E-34 | -33.631 |
| Adrenergic receptor antagonist | 30 | -1.61 | [-1.66,-1.55] | -58.08 | 1.46E-31 | -30.836 |
| **HMGCR inhibitor / statin** | **31** | **-1.67** | **[-1.74,-1.61]** | **-53.31** | **2.79E-31** | **-30.554** |
| Src inhibitor | 30 | -1.71 | [-1.77,-1.64] | -55.4 | 5.65E-31 | -30.248 |
| Calcium channel blocker | 24 | -1.66 | [-1.72,-1.61] | -61.46 | 5.13E-27 | -26.29 |
| Tricyclic antidepressant | 26 | -1.7 | [-1.77,-1.63] | -48.88 | 2.45E-26 | -25.611 |
| Estrogen receptor antagonist | 25 | -1.68 | [-1.75,-1.62] | -52.34 | 2.99E-26 | -25.524 |
| VEGFR inhibitor | 50 | -1.63 | [-1.78,-1.47] | -21.1 | 3.04E-26 | -25.517 |

#### Blocking enzymes from the cholesterol pathway prevents the execution of the EMT-MET program

We first confirmed that HMGCR and DHCR7 were downregulated during the EMT process and came back to a control level during the MET process due to release of the EMT-inducing treatment (**Figure S1**). From the candidates identified above, we chose to test the HMGCR inhibitors, which directly target the cholesterol pathway. We selected two statins, Lovastatin and Simvastatin, and applied them on 2-days-old organoids (**Figure 4A**). Even at high concentration of each statin, we did not observe any change in the phenotypical aspect of the microtissues when added alone (**Figure 4B, C** for *Simvastatin*) or in co-treatment with the EMT-inducing cocktail (*Data not shown*) compared to the EMT-inducing factor alone. However, when statins were added after a pre-treatment with the EMT-inducing cocktail, *i*.*e*. during the EMT-MET program (**Figure 4D**), we observed that the treated spheroids maintained a loose EMT-like phenotype instead of reversing to dense spheroids, as those observed with the Baseline situation (**Figure 4E, F**). This result showed that inhibiting the cholesterol pathway after EMT induction prevented the completion of the MET program, in the absence of EMT inducing factors, highlighting the importance of the regulation of the cholesterol pathway during the EMT-MET programs.

**Figure 4:**
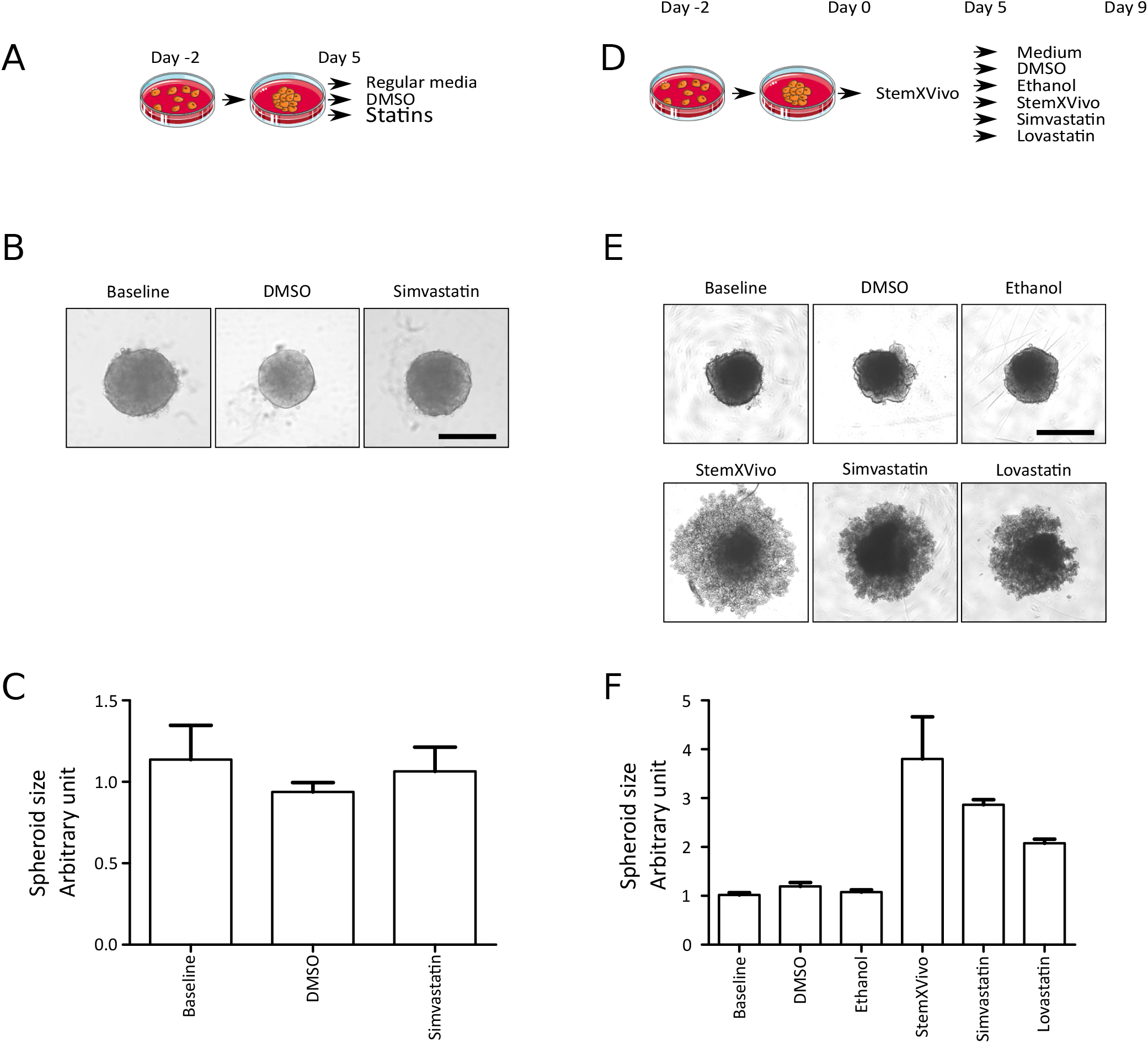
Morphological changes induced with statins depending on the EMT-MET status. For these experiments, cells were grown in ULA plate for 2 days (Day-2) as dense spheroids. Two setting were then used. **A-C)** First setting: From day 0 to day 5, spheroids were either cultured with regular media (baseline), or co-treated with the EMT-inducing cocktail plus DMSO or the EMT-inducing cocktail plus simvastatin. **A)** Experimental setting. **B)** Representative photos of the spheroids in baseline, DMSO and Simvastatin conditions. **C)** Quantification of the spheroid size in arbitrary unit (no significate statistical differences). **D-F)** Second setting: From day 0 to day 5, spheroids were treated with the EMT-inducing cocktail. Then, the EMT-inducing cocktail was washed out and replaced with medium, DMSO (0.5 %), Ethanol (0.5 %), Simvastatin (50 µM), or Lovastatin (50 µM) for 5 days. **D)** Experimental setting. **E)** Representative photos of the spheroids for each condition. **F)** Quantification of the spheroid size, in arbitrary unit. Mean values (+/- SEM) are represented as histograms.

## Discussion

EMT implication in oncogenesis and metastasis has been extensively studied showing the role of many transcription (*e*.*g*. TWIST) and secreted factors (*e*.*g*. TGFβ). However, progress is necessary to better understand this implication and to provide therapeutic targets [9, 22, 23, 38]. The novelty of our study rely on the combination of the broad induction of EMT using an EMT-inducing cocktail, a 3D model and the timeline approach of EMT-MET induction programs.

This allowed the identification of two sets of genes reversibly regulated during either the induction of EMT or the EMT-MET program. The two key pathways retained for our model comprised genes from the EMT pathway, as expected, but also more surprisingly, genes from the cholesterol pathway. When combined as a multigene model, they showed an independent prognostic value in a large dataset of patients with primary colorectal cancer, highlighting the clinical relevance of our finding.

When analyzed in detail, we found that the 14 genes related to the EMT pathway and reversibly modified in response to the EMT-inducing cocktail were genes induced by TGFβ (*PMEPA1, CTGF, TAGLN, CYR61* …). They have also been involved in cell migration (*TNC, IL8, TAGLN, FLNA, SERPINE1, MYL9, CYR61, GPC1*…) in various cancers. At least four of them promoted colorectal liver metastasis through enhancement of the EMT program (*THBS1, TAGLN, ITGA2, FLNA*). Several of them code for surface glycoproteins or integrin-related molecules (*PMEPA1, TAGLN, SERPINE1, THBS1, GPC1*). Some or a combination of these genes might be used as biomarkers for EMT to identify CRC patients potentially more at risk to relapse [39-46].

The involvement of the cholesterol pathway inhibition (during EMT) and re-activation (during EMT-MET) is a new and intriguing finding. The 21 genes of the cholesterol metagene code for most (if not all) enzymes responsible for the synthesis of cholesterol (highlighted in red in **Figure 5**). Cholesterol is essential for cell function and viability. It is a component of the plasma membrane and lipid rafts, a signaling molecule (ligand for estrogen-related receptor alpha (ESRRA)), and a precursor for steroid hormones, Vitamin D and bile acids. The bile acid pathway was also found downregulated during EMT and activated during the EMT-MET programs but did not have prognostic value in our study. Many reports have suggested that cholesterol pathway activation is important for stem cells maintenance in tissues but also for other pro-metastatic function, such as proliferation, survival and resistance to treatment [47]. All these features are associated with pejorative prognosis. Cholesterol synthesis is enhanced in cancer cells compared to normal cells [48, 49]. Additionally, patients with high cholesterol levels have an increased risk for many cancers [48, 49]. But thus far, clinical trials with HMGCR inhibitors (statins) have had mixed results [50, 51]. In our case, activation of the cholesterol synthesis pathway tended to be of better prognosis. Due to its multiple biological roles including in tissue homeostasis, cholesterol might have several roles depending on conditions, cells type or timing. Changes in one of these variables might reveal different functionalities. Compared to previous results, our dynamic model enriched the “EMT field” by highlighting the role of cholesterol regulation in relationship with the triggering of cell detachment signal and cell plasticity. Phenotypic plasticity may provide cancer cells with increased adaptability and resistance, enabling them to respond to a variety of external cues and physiological stresses, including EMT signals. In addition to highlighting the reversibility of the cholesterol pathway activation during EMT-MET programs, we also showed that *in vitro* blockade of the cholesterol pathway altered the execution of the EMT-MET program. One hypothesis is that, in the context of tumor cell motility and migration, EMT requires lowering the synthesis of cholesterol to ensure the destabilization of lipid rafts and/or to allow more flexibility in cell membrane for migration. In this line, in breast and hepatocellular carcinomas, CD44 affinity for cholesterol-rich lipid rafts and retention inside lipid rafts prevent migration [52-56]. Also, statins induce disruption of lipid rafts, impairing tumor cell adhesion, but enhance invasive and metastatic ability of cancer cells [52-56]. The molecular mechanism involved remains however to be fully elucidated. Altogether, these results suggest that beside or through their lipid lowering property, factors such as statins might also influence complex cell biological programs, during oncogenesis such as EMT.

**Figure 5:**
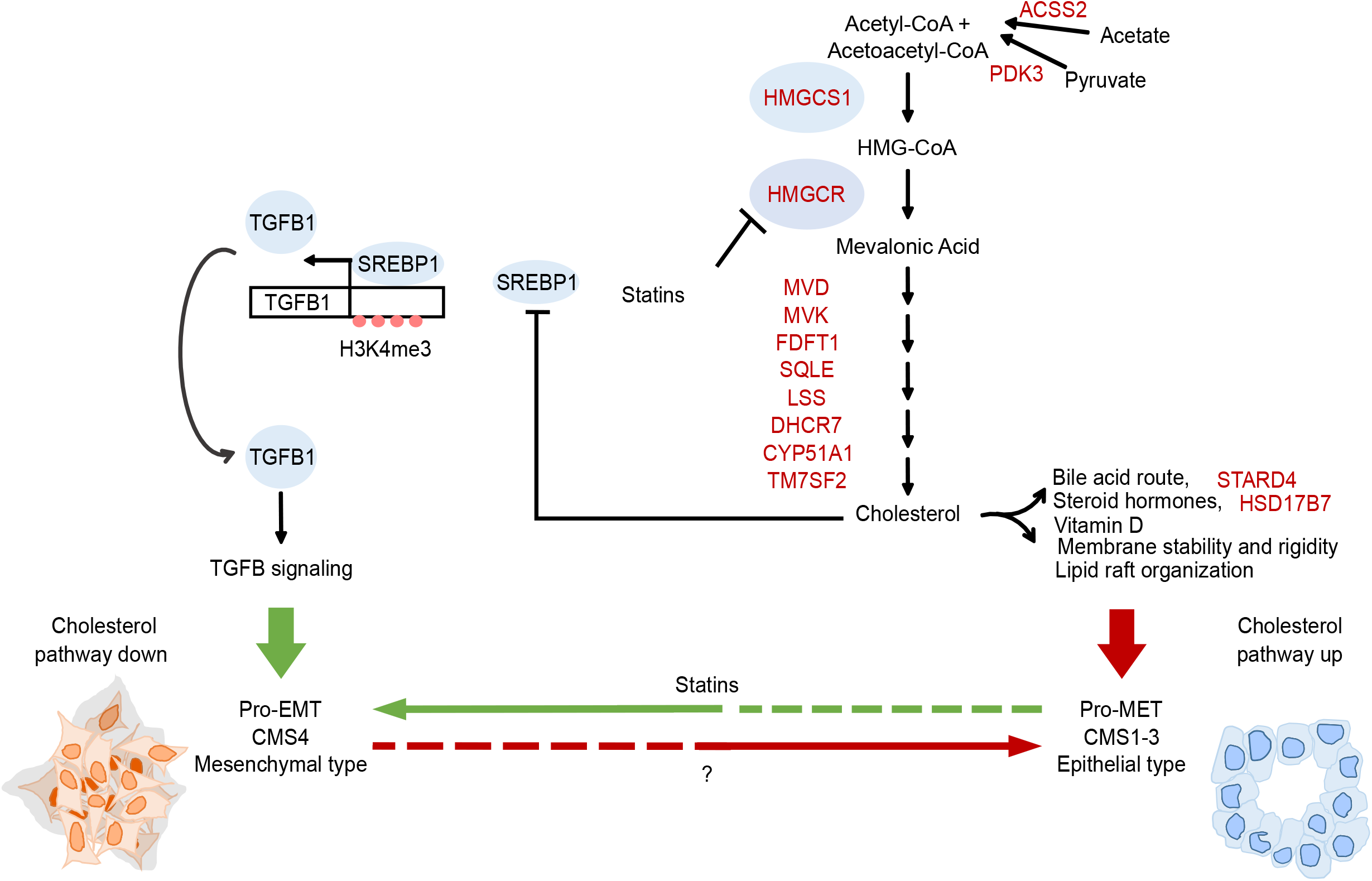
Graphical summary of the potential relationship between EMT activation and the cholesterol pathway. The mevalonate pathway leads to cholesterol synthesis. Cholesterol is involved in multiple biological homeostatic mechanisms (Bile acid, steroid hormones and vitamin D precursor, Membrane stability and rigidity, Lipid raft organization …). Cholesterol inhibits the synthesis of SREBP1, a transcription factor for the TGFβ1 signaling. TGFβ1 signaling is a key feature of the CMS4 mesenchymal class. Our data suggest that low cholesterol in a pro-EMT environment might favor the differentiation of CMS4 mesenchymal tumors, while high cholesterol favored more epithelial tumors. Genes down regulated in our Cholesterol Homeostasis metagene are written in red. Statins are inhibitors of the HMGCR enzyme.

Similar findings (simultaneous opposite involvement of EMT and cholesterol pathways) in pancreatic ductal adenocarcinoma (PDAC) revealed that cholesterol pathway inhibition induces TGFβ signaling and promotes basal differentiation [21]. More specifically, the disruption of cholesterol biosynthesis by *Nsdhl* knockout (in mice) or treatment with statins (in mice and in humans) induces the switch from a glandular to a squamous PDAC subtype (mesenchymal subtype). In line with this, a previous study suggested that human tumor cells belonging to the quasi-mesenchymal/squamous/ basal subtype of PDAC utilize transcriptionally-dominated programs to lose their epithelial phenotype during EMT, while those belonging to the classical/pancreatic progenitor/ADEX subtype rely on protein re-localization to lose their epithelial phenotype during EMT [57]. This recent discovery now reveals that this glandular to squamous PDAC switch can occurs *via* the activation of SREBP1, which induces TGFβ1 expression, autocrine TGFβ-SMAD2/3 signaling and EMT. In line with this major finding, we showed that inducing EMT in our model can switch a cell line from a CMS3 subtype to a CMS4 mesenchymal subtype using a transcriptionally-dominated program [36], connecting this switch to a more pejorative prognosis. Accordingly, we found that the CMS4 subtype [36] was enriched in the “high-risk” prognostic group, *i*.*e* “EMT high, Cholesterol down”. It is thus possible that a general mechanism promoting basal/squamous/mesenchymal differentiation with the transcriptional downregulation of the cholesterol pathway and the upregulation of TGFβ related genes occurs in multiple tumor types (**Figure 5**). Moreover, it was reported that TGFβ-induced mitotic defects in proliferating cells are reversible upon its withdrawal, whereas the acquired genomic abnormalities persist, leading to increased tumorigenic phenotypes. Tumor cells differentiation might thus result from a different degree of EMT depending on several factors, genetic and non-genetic such as the microenvironment and the various treatments [58].

In parallel, and in line with this hypothesis, it would be of interest to identify patients who received statins prior to their diagnosis of colorectal cancer and test if these patient’s tumors are more of the CMS4 subtype. As shown in the PDAC study, this would suggest that the lowering of the cholesterol pathway can indeed influence the subtype of the growing carcinoma, which would be a major discovery in the field. Our *in vitro* data however suggested that cholesterol lowering effects, notably when using statins, occurs only during a specific window to antagonize the MET program. We cannot exclude that a feedback loop between EMT and cholesterol pathways might also exist, which will add a new layer of complexity to this already intricate subject.

## Data Availability

We built a large database gathering our own data set and 10 publicly available sets as described. These latter had been collected from the National Center for Biotechnology Information (NCBI)/Genbank GEO, ArrayExpress and TCGA databases

## Acknowledgements

This work has been supported by Inserm/Institut Paoli-Calmettes and grants from the AP-HM AORC Junior 2018, Canceropole PACA 2018, GIRCI Mediterraneée 2019. A.A. is supported by a postdoctoral fellowship from the “Fondation ARC” (n°PDF20180507565) and the “Fondation de France” (n°00107936). We want to thanks C. Lachaud for sharing the chemidoc equiment.

## Figure legends

**Supplementary Figure 1: Cholesterol homeostasis regulator HMGCR and DHCR7 are down regulated by the EMT-inducing cocktailtreatment**.

Cells were grown in 3D in ULA plate for 2 days (Day-2). At day 0, the EMT and EMT-MET samples were treated with the EMT-inducing cocktail for 5 days. For EMT-MET samples, the spheroids were washed off the EMT-inducing cocktail and grown for 4 more days in normal medium.Cells are collected and lysed withj RIPA buffer before to be subjected to a Western Blot analysis. FIBRONECTINE (FIBRO.) is a mesenchymal marker, CYTOKERATINES (pan CTK) are mostly epithelial markers, both to verify EMT induction by the EMT-inducing cocktail. DHCR7 and HMGCR are regulators of the cholesterol homeostasis pathway. Actin is the loading control.

