## Supplementary figures and images for "Epithelial-to-mesenchymal transition lowers the cholesterol pathway, which influences colon tumors differentiation"

### Supplemental Figure 1

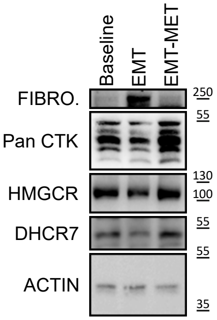
