## Supplemental Table 1 for "Epithelial-to-mesenchymal transition lowers the cholesterol pathway, which influences colon tumors differentiation"

**Table S1:**
List of antibodies used in the study

| Antigen | Host | Manufacturer | CAT # | Dilution |
|---|---|---|---|---|
| *Primary antobodies* | | | | |
| ACTIN | mouse | cell signaling | 3700S | 1/10 000 |
| pan-CYTOKERATIN | rabbit | Abcam | ab9377 | 1/1 000 |
| DHCR7 | rabbit | Abcam | ab103296 | 1/1 000 |
| FIBRONECTIN | sheep | R&D | AF1918 | 1/500 |
| HMGCR | rabbit | Abcam | ab174830 | 1/1 000 |
| OCCLUDIN | mouse | Santa Cruz | sc133256 | 1/1 000 |
| VIMENTIN | chicken | Novus | NB300-223 | 1/1 000 |
| ZEB-1 | rabbit | Abcam | ab203829 | 1/500 |
| *Secondary antobodies* | | | | |
| anti-chicken | donkey | Jackson Immunoresearch | 703-035-155 | 1/10 000 |
| anti-mouse | sheep | SIGMA | NA931 | 1/10 000 |
| anti-rabbit | sheep | SIGMA | NA934 | 1/ 1000 |
| anti-sheep | donkey | Jackson Immunoresearch | 713-035-147 | 1/10 000 |
