## Supplemental Table 2 for "Epithelial-to-mesenchymal transition lowers the cholesterol pathway, which influences colon tumors differentiation"

**Supplementary Table 2 :**
Description of the 11 colon cancer gene expression data sets gathered

| Reference | Source of data | Technological platform | N° of probe sets/genes | N° of colon samples | Primary colon cancer samples (N) |
|---|---|---|---|---|---|
| Jorissen et al., Clin Cancer Res 2009 | GEO database, GSE14333 | Affymetrix, array U133 Plus 2.0 | 54K | 251 | 251 |
| Sheffer et al., Proc Natl Acad Sci 2009 | GEO database, GSE41258 | Affymetrix, array U133 A | 22K | 289 | 168 |
| Staub et al., J Mol Med (Berl) 2009 | GEO database, GSE12945 | Affymetrix, array U133 Plus 2.0 | 54K | 29 | 29 |
| Smith et al., Gastroenterology 2010 | GEO database, GSE17538 | Affymetrix, array U133 Plus 2.0 | 54K | 232 | 232 |
| de Sousa et al., Cell Stem Cell 2011 | GEO database, GSE33113 | Affymetrix, array U133 Plus 2.0 | 54K | 90 | 90 |
| Kenned et al., J Clin Oncol 2011 | Array-Express database, E-MTAB-863 | Affymetrix custom array, ADXCRCG2a520319 | 62K | 215 | 215 |
| Sveen et al., Genome Med 2011 | GEO database, GSE24551 | Affymetrix, Human Exon 1.0 ST array | 22K | 160 | 160 |
| Laibe et al., OMICS 2012 | GEO database, GSE37892 | Affymetrix, array U133 Plus 2.0 | 54K | 130 | 130 |
| Marisa et al., PLoS Med 2013 | GEO database, GSE39582 | Affymetrix, array U133 Plus 2.0 | 54K | 455 | 455 |
| TCGA, COAD | TCGA portal, https://tcga-data.nci.nih.gov | Illumina, RNA sequencing V2 | 25K | 500 | 459 |
| IPC | ---- submission to ArrayExpress ongoing ---- | Affymetrix, array U133 Plus 2.0 | 54K | 50 | 50 |
| | Total | | | 2.401 | 2.239 |
