## Supplemental Table 3 for "Epithelial-to-mesenchymal transition lowers the cholesterol pathway, which influences colon tumors differentiation"

**Supplementary Table 3:**
DAVID ontology analysis of the clusters 1 and 2 using GO Biological processes (GO:BP) database

| Clusters | GO:BP term | Count | % | P-value | Genes |
|---|---|---|---|---|---|
| luster 1 | GO:0007155~cell adhesion | 14 | 7.57 | 7.87E-05 | NRP2, ITGA2, TNC, NEDD9, HAPLN3, THBS1, CYR61, EFNB2, KITLG, ALCAM, PODXL, ZYX, ITGB8, VCL |
| | GO:0030335~positive regulation of cell migration | 9 | 4.86 | 1.15E-04 | SEMA7A, EDN1, PODXL, PDGFB, PDGFA, THBS1, F3, CYR61, CCAR1 |
| | GO:0048661~positive regulation of smooth muscle cell proliferation | 6 | 3.24 | 1.15E-04 | EDN1, ITGA2, PDGFB, IL18, PTGS2, THBS1 |
| | GO:0030198~extracellular matrix organization | 9 | 4.86 | 1.78E-04 | ITGA2, SERPINE1, PDGFB, DAG1, TNC, ITGB8, PDGFA, THBS1, CYR61 |
| | GO:0007507~heart development | 8 | 4.32 | 6.68E-04 | ADAM19, EDN1, NRP2, FLRT3, OSR1, PDGFB, PCSK5, PPARD |
| | GO:0002576~platelet degranulation | 6 | 3.24 | 1.41E-03 | SERPINE1, PDGFB, PDGFA, FLNA, THBS1, VCL |
| | GO:0050919~negative chemotaxis | 4 | 2.16 | 2.48E-03 | SEMA7A, NRP2, FLRT3, PDGFA |
| | GO:0042060~wound healing | 5 | 2.70 | 3.84E-03 | TNC, PDGFA, ELK3, PPARD, DCBLD2 |
| | GO:0030036~actin cytoskeleton organization | 6 | 3.24 | 3.87E-03 | GAS2L3, FGD6, MKL1, PDGFA, ARHGEF18, SSH2 |
| | GO:0045184~establishment of protein localization | 4 | 2.16 | 3.96E-03 | ITGA2, MDM2, FLNA, LIMS1 |
| | GO:0007267~cell-cell signaling | 8 | 4.32 | 4.30E-03 | EFNB2, EDN1, IL18, ZYX, PDGFA, NMUR2, PCSK5, CYR61 |
| | GO:0010595~positive regulation of endothelial cell migration | 4 | 2.16 | 5.87E-03 | EDN1, NRP2, ETS1, THBS1 |
| | GO:0044849~estrous cycle | 3 | 1.62 | 7.95E-03 | MMP7, ANXA1, ETS1 |
| | GO:0007229~integrin-mediated signaling pathway | 5 | 2.70 | 8.14E-03 | SEMA7A, ITGA2, ZYX, ITGB8, NEDD9 |
| | GO:0042493~response to drug | 8 | 4.32 | 1.11E-02 | FOSL1, ANXA1, ITGA2, MDM2, NFATC2, GCLM, PTGS2, THBS1 |
| | GO:0043406~positive regulation of MAP kinase activity | 4 | 2.16 | 1.17E-02 | EDN1, KITLG, PDGFB, PDGFA |
| | GO:0009612~response to mechanical stimulus | 4 | 2.16 | 1.17E-02 | FOSL1, TNC, ETS1, THBS1 |
| | GO:0045987~positive regulation of smooth muscle contraction | 3 | 1.62 | 1.20E-02 | EDN1, ITGA2, PTGS2 |
| | GO:0008284~positive regulation of cell proliferation | 10 | 5.41 | 1.25E-02 | EFNB2, FOSL1, EDN1, MDM2, PDGFB, TNC, PDGFA, ETS1, THBS1, CCAR1 |
| | GO:0050727~regulation of inflammatory response | 4 | 2.16 | 1.39E-02 | SEMA7A, ANXA1, ZYX, PTGS2 |
| | GO:0014068~positive regulation of phosphatidylinositol 3-kinase signaling | 4 | 2.16 | 1.51E-02 | PDGFB, IL18, PDGFA, PPARD |
| | GO:0032877~positive regulation of DNA endoreduplication | 2 | 1.08 | 1.59E-02 | E2F7, E2F8 |

| | | | | | |
|---|---|---|---|---|---|
| c | GO:0010512~negative regulation of phosphatidylinositol biosynthetic process | 2 | 1.08 | 1.59E-02 | PDGFB, PDGFA |
| | GO:0061614~pri-miRNA transcription from RNA polymerase II promoter | 2 | 1.08 | 1.59E-02 | FOSL1, ETS1 |
| | GO:0002040~sprouting angiogenesis | 3 | 1.62 | 1.68E-02 | THBS1, E2F7, E2F8 |
| | GO:0043154~negative regulation of cysteine-type endopeptidase activity involved in apoptotic process | 4 | 2.16 | 1.78E-02 | TNFAIP8, MKL1, MDM2, THBS1 |
| | GO:0010628~positive regulation of gene expression | 7 | 3.78 | 1.87E-02 | OSR1, MDM2, PDGFB, TNC, ITGB8, LIMS1, PPARD |
| | GO:0050729~positive regulation of inflammatory response | 4 | 2.16 | 2.06E-02 | ITGA2, SERPINE1, IL18, ETS1 |
| | GO:0035793~positive regulation of metanephric mesenchymal cell migration by platelet-derived growth factor receptor-beta signaling pathway | 2 | 1.08 | 2.38E-02 | PDGFB, PDGFA |
| | GO:0046718~viral entry into host cell | 4 | 2.16 | 2.61E-02 | EFNB2, ITGA2, DAG1, CLDN1 |
| | GO:1900026~positive regulation of substrate adhesion-dependent cell spreading | 3 | 1.62 | 2.68E-02 | FLNA, UNC13D, LIMS1 |
| | GO:0051897~positive regulation of protein kinase B signaling | 4 | 2.16 | 2.96E-02 | IL18, PDGFA, THBS1, F3 |
| | GO:0031394~positive regulation of prostaglandin biosynthetic process | 2 | 1.08 | 3.15E-02 | ANXA1, PTGS2 |
| | GO:0035655~interleukin-18-mediated signaling pathway | 2 | 1.08 | 3.15E-02 | PDGFB, IL18 |
| | GO:0001890~placenta development | 3 | 1.62 | 3.17E-02 | GJB3, E2F7, E2F8 |
| | GO:0001525~angiogenesis | 6 | 3.24 | 3.33E-02 | NRP2, SERPINE1, IL18, PDGFA, PTGS2, ELK3 |
| | GO:0007160~cell-matrix adhesion | 4 | 2.16 | 3.53E-02 | ITGA2, ZYX, ITGB8, VCL |
| | GO:0043547~positive regulation of GTPase activity | 10 | 5.41 | 3.73E-02 | TIAM2, KITLG, FGD6, PDGFB, PDGFA, ARHGAP29, ARHGEF18, ARHGAP17, AXIN2, LIMS1 |
| | GO:0060718~chorionic trophoblast cell differentiation | 2 | 1.08 | 3.93E-02 | E2F7, E2F8 |
| | GO:0010757~negative regulation of plasminogen activation | 2 | 1.08 | 3.93E-02 | SERPINE1, THBS1 |
| | GO:0071456~cellular response to hypoxia | 4 | 2.16 | 4.15E-02 | EDN1, MDM2, PTGS2, PPARD |
| | GO:0007566~embryo implantation | 3 | 1.62 | 4.42E-02 | PTGS2, PCSK5, PPARD |
| | GO:0045740~positive regulation of DNA replication | 3 | 1.62 | 4.42E-02 | KITLG, PDGFB, PDGFA |
| | GO:0014823~response to activity | 3 | 1.62 | 4.61E-02 | EDN1, GCLM, PPARD |
| | GO:0006695~cholesterol biosynthetic process | 16 | 6.18 | 4.42E-20 | IDI1, MVK, HMGCS1, INSIG1, CYP51A1, MSMO1, HMGCR, HSD17B7, LSS, TM7SF2, SQLE, EBP, MVD, DHCR7, CFTR, FDFT1 |
| | GO:0008299~isoprenoid biosynthetic process | 6 | 2.32 | 3.24E-07 | IDI1, MVK, HMGCS1, MVD, HMGCR, FDFT1 |
| | GO:0055114~oxidation-reduction process | 21 | 8.11 | 1.31E-05 | MAOA, CYP51A1, MSMO1, HMGCR, FAM213A, HSD17B7, PPOX, PTGR2, TM7SF2, SQLE, PRDX5, ALDH6A1, P4HA1, RDH11, SCD, HSD17B2, CYP1B1, CRYL1, DHCR7, STEAP2, FDFT1 |

| | | | | | |
|---|---|---|---|---|---|
| Cluster 2 | GO:0008203~cholesterol metabolic process | 6 | 2.32 | 1.03E-03 | LIPE, SQLE, EBP, INSIG1, LRP5, NR0B2 |
| | GO:0002053~positive regulation of mesenchymal cell proliferation | 4 | 1.54 | 3.06E-03 | FGF9, LRP5, IRS2, FGFR2 |
| | GO:0016126~sterol biosynthetic process | 3 | 1.16 | 5.40E-03 | SQLE, MSMO1, TM7SF2 |
| | GO:0006694~steroid biosynthetic process | 4 | 1.54 | 5.56E-03 | CYP51A1, HSD17B2, LSS, FDFT1 |
| | GO:0010510~regulation of acetyl-CoA biosynthetic process from pyruvate | 3 | 1.16 | 7.80E-03 | PDK4, PDK3, PDK1 |
| | GO:0008610~lipid biosynthetic process | 3 | 1.16 | 1.06E-02 | ACSS2, ACSL1, FDFT1 |
| | GO:0014066~regulation of phosphatidylinositol 3-kinase signaling | 5 | 1.93 | 1.18E-02 | ERBB3, FGF9, IRS2, PPP2R5C, FGFR2 |
| | GO:0008631~intrinsic apoptotic signaling pathway in response to oxidative stress | 3 | 1.16 | 1.21E-02 | CYP1B1, MAP3K5, PDK1 |
| | GO:0008152~metabolic process | 7 | 2.70 | 1.22E-02 | LIPE, ACSS2, ACSL1, INSIG1, ECI2, ACSS1, ENPP5 |
| | GO:0042472~inner ear morphogenesis | 4 | 1.54 | 2.10E-02 | FGF9, INSIG1, FGFR2, ATOH1 |
| | GO:0019542~propionate biosynthetic process | 2 | 0.77 | 2.25E-02 | ACSS2, ACSS1 |
| | GO:0019413~acetate biosynthetic process | 2 | 0.77 | 2.25E-02 | ACSS2, ACSS1 |
| | GO:0019427~acetyl-CoA biosynthetic process from acetate | 2 | 0.77 | 2.25E-02 | ACSS2, ACSS1 |
| | GO:0016125~sterol metabolic process | 3 | 1.16 | 2.32E-02 | EBP, CYP51A1, CYP1B1 |
| | GO:0009235~cobalamin metabolic process | 3 | 1.16 | 2.32E-02 | LMBRD1, TCN1, MMAB |
| | GO:0010906~regulation of glucose metabolic process | 3 | 1.16 | 2.54E-02 | PDK4, PDK3, PDK1 |
| | GO:0050679~positive regulation of epithelial cell proliferation | 4 | 1.54 | 3.05E-02 | FGF9, HYAL1, GLUL, FGFR2 |
| | GO:0050746~regulation of lipoprotein metabolic process | 2 | 0.77 | 3.36E-02 | DGAT2, LIPG |
| | GO:0006725~cellular aromatic compound metabolic process | 2 | 0.77 | 3.36E-02 | SQLE, CYP1B1 |
| | GO:0019287~isopentenyl diphosphate biosynthetic process, mevalonate pathway | 2 | 0.77 | 3.36E-02 | MVK, MVD |
| | GO:0006006~glucose metabolic process | 4 | 1.54 | 4.03E-02 | PDK4, PDK3, IRS2, PDK1 |
| | GO:0061304~retinal blood vessel morphogenesis | 2 | 0.77 | 4.45E-02 | LRP5, CYP1B1 |
| | GO:0097411~hypoxia-inducible factor-1alpha signaling pathway | 2 | 0.77 | 4.45E-02 | PDK3, PDK1 |
| | GO:0033489~cholesterol biosynthetic process via desmosterol | 2 | 0.77 | 4.45E-02 | EBP, DHCR7 |
| | GO:0009298~GDP-mannose biosynthetic process | 2 | 0.77 | 4.45E-02 | PMM1, MPI |
| | GO:0033490~cholesterol biosynthetic process via lathosterol | 2 | 0.77 | 4.45E-02 | EBP, DHCR7 |
| | GO:0042572~retinol metabolic process | 3 | 1.16 | 4.50E-02 | DGAT2, RDH11, CYP1B1 |
