## Supplemental Table 4 for "Epithelial-to-mesenchymal transition lowers the cholesterol pathway, which influences colon tumors differentiation"

**Suplementary Table 5 :**
Comparison of prognostic information of metagene models based on unique and combined variables in the public colon cancer database (N=1,837)

| **Cox, models comparison Likelihood ($X^2$ Chi-squared)** | | |
|---|---|---|
| EMT | $LRX^2$ | 11.81 |
| | p-value | 5.89E-04 |
| CHL | $LRX^2$ | 5.80 |
| | p-value | 1.61E-02 |
| EMT+CHL | $LRX^2$ | 17.91 |
| | p-value | 1.29E-04 |
| **EMT** | **$DLRX^2$** | **6.10** |
| **vs. EMT+CHL** | **p-value** | **1.35E-02** |
| **CHL** | **$DLRX^2$** | **12.12** |
| **vs. EMT+CHL** | **p-value** | **5.00E-04** |
