## Supplemental Table 5 for "Epithelial-to-mesenchymal transition lowers the cholesterol pathway, which influences colon tumors differentiation"

**Supplementary Table 6:**
Connectivity map (L1000), 14 EMT vs. 21g CHL GSEA core genes

| Drug class (moa) | pertubagen | Number of sigificant | Normalised connectivity score | ncs average 95% CI | Reproducibility of ncs per | |
|---|---|---|---|---|---|---|
| | | | | | t | p.value |
| Dopamine receptor antagonist | clozapine,flupentixol,sertindole,fluphenazine,thiothixene,prochlorperazine,asenapine,promazine,thioridazine,fluspirilene,spiperone,trifluoperazine,butaclamol,iloperidone,nemonapride,thioproperazine,amperozide,pimozide,mesoridazine,spiramide,perphenazine,triflupromazine,cariprazine,acepromazine,isofloxythepin,haloperidol,brexpiprazole | 93 | -1.64 | [-1.67,-1.61] | -98.47 | 4.83E-95 |
| EGFR inhibitor | golvatinib,lapatinib,gefitinib,neratinib,sunitinib,vandetanib,tivozanib,pazopanib,regorafenib,cediranib,afatinib,canertinib,foretinib,nintedanib,dovitinib,dacomitinib,pelitinib | 80 | -1.65 | [-1.74,-1.55] | -33.31 | 2.86E-48 |
| Serotonin receptor antagonist | clozapine,ketanserin,vilazodone,sertindole,bemesetron,idalopirdine,metergoline,asenapine,sertraline,vortioxetine,clomipramine,iloperidone,spiramide,amitriptyline,pimavanserin,norcyclobenzaprine,pizotifen | 44 | -1.63 | [-1.68,-1.58] | -62.43 | 7.92E-44 |
| PDGFR inhibitor | tandutinib,masitinib,sunitinib,dasatinib,pazopanib,ponatinib,imatinib,regorafenib,nintedanib,dovitinib | 37 | -1.65 | [-1.71,-1.59] | -57.98 | 3.75E-37 |
| Bcr-Abl inhibitor | dasatinib,ponatinib,imatinib,bosutinib,flumatinib,tozasertib,nilotinib | 33 | -1.67 | [-1.72,-1.62] | -63.64 | 2.85E-35 |
| Abl inhibitor | dasatinib,ponatinib,imatinib,bosutinib,flumatinib,tozasertib,nilotinib | 33 | -1.67 | [-1.72,-1.62] | -63.64 | 2.85E-35 |
| Histamine receptor antagonist | DPPE,cyproheptadine,doxepin,methapyrilene,clemastine,astemizole,loratadine,promethazine,thioperamide,terfenadine,levomequitazine,tecastemizole,dexchlorpheniramine,desloratadine,iodophenpropit,zolantidine,azelastine | 33 | -1.61 | [-1.66,-1.56] | -62.66 | 4.67E-35 |
| KIT inhibitor | tandutinib,masitinib,sunitinib,dasatinib,pazopanib,imatinib,regorafenib,cediranib,midostaurin | 34 | -1.66 | [-1.72,-1.6] | -56.25 | 2.34E-34 |
| Adrenergic receptor antagonist | talinolol,nicergoline,morphothebaine,prazosin,phentolamine,carvedilol,sotalol,suloctidil,nebivolol,cyclazosin,dronedarone,propranolol,niguldipine | 30 | -1.61 | [-1.66,-1.55] | -58.08 | 1.46E-31 |

| | | | | | | |
|---|---|---|---|---|---|---|
| HMGCR inhibitor / statin | mevastatin,fluvastatin,rosuvastatin,atorvastatin,pravastatin,pitavastatin,simvastatin,cerivastatin,lovastatin | 31 | -1.67 | [-1.74,-1.61] | -53.31 | 2.79E-31 |
| Src inhibitor | masitinib,dasatinib,saracatinib,bosutinib | 30 | -1.71 | [-1.77,-1.64] | -55.4 | 5.65E-31 |
| Calcium channel blocker | mibefradil,penfluridol,lidoflazine,amlodipine,lomerizine,verapamil,niguldipine,ionomycin,tetrandrine | 24 | -1.66 | [-1.72,-1.61] | -61.46 | 5.13E-27 |
| Tricyclic antidepressant | trimipramine,maprotiline,nortriptyline,protriptyline,desipramine | 26 | -1.7 | [-1.77,-1.63] | -48.88 | 2.45E-26 |
| Estrogen receptor antagonist | tamoxifen,toremifene,fulvestrant,raloxifene,clomifene | 25 | -1.68 | [-1.75,-1.62] | -52.34 | 2.99E-26 |
| VEGFR inhibitor | golvatinib,sunitinib,vandetanib,tivozanib,pazopanib,regorafenib,cediranib,foretinib,nintedanib,dovitinib | 50 | -1.63 | [-1.78,-1.47] | -21.1 | 3.04E-26 |
| FLT3 inhibitor | tandutinib,sunitinib,ponatinib,tozasertib,dovitinib,midostaurin,quizartinib | 25 | -1.7 | [-1.77,-1.63] | -50.81 | 6.03E-26 |
| Norepinephrine inhibitor | indatraline,trimipramine,maprotiline,amoxapine,imipramine,duloxetine,amitriptyline | 22 | -1.65 | [-1.72,-1.57] | -45.34 | 1.95E-22 |
| Selective estrogen receptor modulator | bazedoxifene,toremifene,tamoxifen,raloxifene | 17 | -1.65 | [-1.72,-1.58] | -49.01 | 7.24E-19 |
| Adrenergic receptor agonist | salmeterol,buphenine,indacaterol,isoxsuprine,cyclobenzaprine,norcyclobenzaprine | 11 | -1.58 | [-1.65,-1.51] | -47.74 | 3.92E-13 |
| Opioid receptor agonist | FIT,loperamide | 13 | -1.77 | [-1.89,-1.65] | -32.4 | 4.72E-13 |
| Sterol demethylase inhibitor | ketoconazole,tioconazole,sulconazole,econazole,sertaconazole,terconazole | 12 | -1.66 | [-1.76,-1.56] | -37.09 | 6.60E-13 |
| FGFR inhibitor | danusertib,regorafenib,nintedanib,dovitinib | 12 | -1.7 | [-1.82,-1.59] | -33.05 | 2.33E-12 |
| HDAC inhibitor | panobinostat,dacinostat,apicidin,ISOX,scriptaid,droxinostat,resminostat,givinostat | 11 | -1.52 | [-1.6,-1.43] | -39.06 | 2.89E-12 |
| Aurora kinase inhibitor | danusertib,tozasertib | 11 | -1.75 | [-1.86,-1.65] | -36.68 | 5.39E-12 |
| RET tyrosine kinase inhibitor | vandetanib | 11 | -1.68 | [-1.81,-1.55] | -28.66 | 6.22E-11 |
| Potassium channel antagonist | amiodarone,linopirdine,paxilline,ibutilide | 7 | -1.55 | [-1.6,-1.51] | -79.05 | 2.76E-10 |
| JAK inhibitor | ruxolitinib,tozasertib | 9 | -1.76 | [-1.89,-1.63] | -31.67 | 1.08E-09 |
| ALK inhibitor | DMH1,crizotinib,ceritinib | 7 | -1.55 | [-1.61,-1.48] | -56.04 | 2.17E-09 |
| Monoamine oxidase inhibitor | pirlindole,tetrindole | 9 | -1.64 | [-1.8,-1.49] | -24.01 | 9.65E-09 |

| | | | | | | |
|---|---|---|---|---|---|---|
| Bacterial permeability inducer | lasalocid | 8 | -1.65 | [-1.78,-1.53] | -30.78 | 9.87E-09 |
| BTK inhibitor | acalabrutinib,ibrutinib | 7 | -1.58 | [-1.67,-1.49] | -42.4 | 1.15E-08 |
| Bacterial cell wall synthesis inhibitor | fenticonazole,miconazole,dicloxacillin,econazole | 6 | -1.52 | [-1.59,-1.46] | -62.76 | 1.94E-08 |
| Dopamine receptor agonist | rotigotine,metergoline | 8 | -1.65 | [-1.79,-1.51] | -27.58 | 2.12E-08 |
| Sigma receptor antagonist | rimcazole,siramesine | 7 | -1.66 | [-1.78,-1.54] | -34.98 | 3.64E-08 |
| Calmodulin inhibitor | dexniguldipine,berbamine,zaldaride | 8 | -1.72 | [-1.89,-1.56] | -25.06 | 4.11E-08 |
| PKC inhibitor | enzastaurin,midostaurin,sotrastaurin | 7 | -1.65 | [-1.78,-1.53] | -32.31 | 5.85E-08 |
| Cyclooxygenase inhibitor | phenazone,ibuprofen,phenacetin,diclofenac,pterostilbene,indome tacin | 6 | -1.51 | [-1.59,-1.43] | -48.81 | 6.82E-08 |
| Adenosine receptor antagonist | dilazep,mefloquine | 7 | -1.66 | [-1.8,-1.52] | -28.44 | 1.25E-07 |
| Protein synthesis inhibitor | retapamulin,puromycin,rifabutin,thiostrepton | 5 | -1.58 | [-1.65,-1.51] | -63.96 | 3.58E-07 |
| Serotonin receptor agonist | vortioxetine,cyclobenzaprine | 6 | -1.66 | [-1.78,-1.54] | -34.5 | 3.85E-07 |
| Norepinephrine reuptake inhibitor | maprotiline,amitriptyline | 7 | -1.78 | [-1.96,-1.59] | -23.24 | 4.16E-07 |
| Serotonin reuptake inhibitor | imipramine,duloxetine,amitriptyline | 7 | -1.71 | [-1.89,-1.53] | -23.08 | 4.34E-07 |
| P-glycoprotein inhibitor | zosuquidar,elacridar | 6 | -1.6 | [-1.73,-1.46] | -30.65 | 6.94E-07 |
| Selective serotonin reuptake inhibitor | fluvoxamine,fluoxetine,paroxetine | 7 | -1.72 | [-1.93,-1.51] | -20.3 | 9.29E-07 |
| RAF inhibitor | dabrafenib,regorafenib | 6 | -1.6 | [-1.75,-1.45] | -26.9 | 1.33E-06 |
| Acetylcholine receptor antagonist | piperidolate,pentoxyverine,trihexyphenidyl,mebeverine,metixene | 6 | -1.62 | [-1.77,-1.46] | -26.78 | 1.36E-06 |
| DNA inhibitor | enrofloxacin,edoxudine,butylparaben,niclosamide,clofazimine | 6 | -1.63 | [-1.79,-1.48] | -26.57 | 1.41E-06 |
| Ephrin inhibitor | dasatinib | 5 | -1.62 | [-1.79,-1.46] | -27.02 | 1.12E-05 |
| Tyrosine kinase inhibitor | dasatinib | 5 | -1.62 | [-1.79,-1.46] | -27.02 | 1.12E-05 |
| Carnitine palmitoyltransferase inhibitor | perhexiline | 5 | -1.63 | [-1.87,-1.4] | -19.49 | 4.08E-05 |
| MEK inhibitor | trametinib,cobimetinib,selumetinib | 5 | -1.61 | [-1.85,-1.38] | -19.28 | 4.27E-05 |

| | | | | | | |
|---|---|---|---|---|---|---|
| Sodium channel inhibitor | dichlorobenzamil | 5 | -1.59 | [-1.83,-1.36] | -18.91 | 4.60E-05 |
| RET inhibitor | sunitinib,regorafenib | 5 | -1.68 | [-1.94,-1.43] | -18.63 | 4.89E-05 |
| Cytochrome P450 inhibitor | arbidol,androstenedione,clotrimazole | 4 | -1.57 | [-1.74,-1.4] | -29.11 | 8.90E-05 |
| HSP inhibitor | radicicol,alvespimycin | 4 | -1.54 | [-1.72,-1.36] | -26.92 | 1.12E-04 |
| Kinesin inhibitor | ispinesib | 4 | -1.66 | [-1.86,-1.46] | -26.6 | 1.17E-04 |
| Histamine receptor agonist | alimemazine,amodiaquine | 5 | -1.69 | [-2.02,-1.35] | -13.77 | 1.61E-04 |
| Carcinogen | benzofuran,rhodomyrtoxin | 3 | -1.61 | [-1.71,-1.51] | -71.39 | 1.96E-04 |
| HIV inhibitor | ritonavir,saquinavir | 4 | -1.61 | [-1.85,-1.38] | -21.52 | 2.19E-04 |
| Phosphodiesterase inhibitor | mirodenafil,roflumilast,ibudilast | 3 | -1.48 | [-1.59,-1.37] | -60.28 | 2.75E-04 |
| Bacterial 30S ribosomal subunit inhibitor | eperezolid,spectinomycin,minocycline | 3 | -1.54 | [-1.65,-1.42] | -58.01 | 2.97E-04 |
| Cannabinoid receptor agonist | lylamine | 4 | -1.6 | [-1.86,-1.34] | -19.37 | 3.01E-04 |
| ERBB2 inhibitor | lapatinib | 4 | -1.72 | [-2.05,-1.39] | -16.4 | 4.93E-04 |
| Acetylcholine receptor agonist | bifemelane,desmethylclozapine | 4 | -1.7 | [-2.05,-1.35] | -15.36 | 6.00E-04 |
| PPAR receptor agonist | carbacyclin,rosiglitazone,pterostilbene | 3 | -1.5 | [-1.66,-1.33] | -38.92 | 6.60E-04 |
| Estrogen receptor agonist | estradiol,PPT | 3 | -1.55 | [-1.74,-1.35] | -33.94 | 8.67E-04 |
| ATPase inhibitor | DBeQ,blebbistatin,thapsigargin | 3 | -1.51 | [-1.7,-1.31] | -33.64 | 8.83E-04 |
| Protein synthesis stimulant | dehydroepiandrosterone | 2 | -1.64 | [-1.67,-1.61] | -670.55 | 9.49E-04 |
| Thrombin inhibitor | skatole,voapaxar | 4 | -1.7 | [-2.13,-1.27] | -12.69 | 1.05E-03 |
| BCL inhibitor | obatoclax | 4 | -1.63 | [-2.06,-1.19] | -12.01 | 1.24E-03 |
| NFKB inhibitor | parthenolide,mepacrine,cepharanthine | 4 | -1.77 | [-2.26,-1.28] | -11.51 | 1.41E-03 |
| Nitric oxide production inhibitor | brazilin | 3 | -1.58 | [-1.86,-1.31] | -24.64 | 1.64E-03 |
| Tubulin inhibitor | fenbendazole,epothilone,paclitaxel | 3 | -1.66 | [-2.06,-1.26] | -17.82 | 3.13E-03 |
| Lipase inhibitor | orlistat | 2 | -1.72 | [-1.84,-1.61] | -192.45 | 3.31E-03 |
| Immunosuppressant | fingolimod | 3 | -1.67 | [-2.09,-1.25] | -17.08 | 3.41E-03 |
| Sphingosine 1 phosphate receptor agonist | fingolimod | 3 | -1.67 | [-2.09,-1.25] | -17.08 | 3.41E-03 |

| | | | | | | |
|---|---|---|---|---|---|---|
| Potassium channel activator | retigabine,hexachlorophene | 3 | -1.64 | [-2.09,-1.19] | -15.66 | 4.06E-03 |
| Vesicular monoamine transporter inhibitor | reserpine | 3 | -1.62 | [-2.08,-1.16] | -15.21 | 4.30E-03 |
| Leukotriene inhibitor | quiflapon | 2 | -1.5 | [-1.64,-1.37] | -140.92 | 4.52E-03 |
| HIF Inhibitor | IOX2 | 2 | -1.46 | [-1.63,-1.28] | -104.86 | 6.07E-03 |
| Neural Wiskott-Aldrich syndrome protein inhibitor | wiskostatin | 3 | -1.67 | [-2.24,-1.11] | -12.76 | 6.09E-03 |
| Vasopressin receptor antagonist | tolvaptan,conivaptan | 3 | -1.69 | [-2.26,-1.12] | -12.72 | 6.12E-03 |
| STAT inhibitor | niclosamide | 2 | -1.67 | [-1.98,-1.35] | -67.24 | 9.47E-03 |
| Cholesterol inhibitor | ezetimibe | 2 | -1.58 | [-1.92,-1.25] | -60.54 | 1.05E-02 |
| Niemann-Pick C1-like 1 protein antagonist | ezetimibe | 2 | -1.58 | [-1.92,-1.25] | -60.54 | 1.05E-02 |
| Antifungal | methylparaben,flucytosine | 2 | -1.47 | [-1.82,-1.12] | -53.26 | 1.20E-02 |
| Histone demethylase inhibitor | methylstat | 2 | -1.47 | [-1.84,-1.09] | -49.38 | 1.29E-02 |
| DNA intercalating agent | rhodomyrtoxin | 2 | -1.62 | [-2.04,-1.2] | -48.95 | 1.30E-02 |
| Insulin sensitizer | rosiglitazone,metformin | 2 | -1.5 | [-1.93,-1.07] | -44.33 | 1.44E-02 |
| Opioid receptor antagonist | chloroxine,naltrindole | 2 | -1.47 | [-1.9,-1.05] | -44.34 | 1.44E-02 |
| HCV inhibitor | grazoprevir,dasabuvir | 2 | -1.65 | [-2.16,-1.14] | -41.15 | 1.55E-02 |
| Tachykinin antagonist | netupitant | 2 | -1.78 | [-2.43,-1.12] | -34.3 | 1.86E-02 |
| Glucocorticoid receptor agonist | flumetasone,dexamethasone | 2 | -1.52 | [-2.28,-0.76] | -25.52 | 2.49E-02 |
| Importin inhibitor | importazole | 2 | -1.59 | [-2.43,-0.76] | -24.39 | 2.61E-02 |
| Membrane integrity inhibitor | tetracaine | 2 | -1.67 | [-2.88,-0.45] | -17.48 | 3.64E-02 |
| Ribonucleoside reductase inhibitor | gemcitabine | 2 | -1.55 | [-2.74,-0.36] | -16.52 | 3.85E-02 |
| Microsomal trigylceride transfer protein inhibitor | lomitapide | 2 | -1.83 | [-3.25,-0.4] | -16.31 | 3.90E-02 |
| Antihistamine | homochlorcyclizine | 2 | -1.55 | [-3.01,-0.1] | -13.57 | 4.68E-02 |

| | | | | | | |
|---|---|---|---|---|---|---|
| P38 MAPK inhibitor | doramapimod | 2 | -1.74 | [-3.78,0.3] | -10.87 | 5.84E-02 |
| Butyrylcholinesterase inhibitor | profenamine | 2 | -1.66 | [-3.81,0.49] | -9.79 | 6.48E-02 |
| Cholinergic receptor antagonist | profenamine | 2 | -1.66 | [-3.81,0.49] | -9.79 | 6.48E-02 |
| Bacterial DNA inhibitor | enrofloxacin,clofazimine | 2 | -1.68 | [-4.39,1.04] | -7.83 | 8.09E-02 |
| MTOR inhibitor | sirolimus,everolimus | 2 | 0.16 | [-23.64,23.95] | 0.08 | 9.47E-01 |
| Antimalarial | halofantrine | 1 | -1.85 | NA | NA | NA |
| Aldose reductase inhibitor | isoquercetin | 1 | -1.81 | NA | NA | NA |
| Anesthetic - local | oxetacaine | 1 | -1.73 | NA | NA | NA |
| MDM inhibitor | RITA | 1 | -1.63 | NA | NA | NA |
| Lanosterol demethylase inhibitor | econazole | 1 | -1.59 | NA | NA | NA |
| Imidazoline ligand | clotrimazole | 1 | -1.59 | NA | NA | NA |
| Enzyme inducer | climbazole | 1 | -1.58 | NA | NA | NA |
| Anti-HCVE2 | metitepine | 1 | -1.57 | NA | NA | NA |
| Mitochondrial oxidative phosphorylation uncoupler | FCCP | 1 | -1.57 | NA | NA | NA |
| Immunostimulant | tetramisole | 1 | -1.57 | NA | NA | NA |
| Bacterial antifolate | sulfabenzamide | 1 | -1.57 | NA | NA | NA |
| Fungal 1,3-beta-D-glucan synthase inhibitor | anidulafungin | 1 | -1.54 | NA | NA | NA |
| Smoothened receptor antagonist | sonidegib | 1 | -1.54 | NA | NA | NA |
| Leukotriene receptor antagonist | ibudilast | 1 | -1.53 | NA | NA | NA |
| Prostanoid receptor antagonist | benzydamine | 1 | -1.53 | NA | NA | NA |
| Cytokine production inhibitor | mepacrine | 1 | -1.53 | NA | NA | NA |
| p53 activator | mepacrine | 1 | -1.53 | NA | NA | NA |
| Hemoglobin antagonist | mefloquine | 1 | -1.52 | NA | NA | NA |

| | | | | | | |
|---|---|---|---|---|---|---|
| Sodium/glucose cotransporter inhibitor | dapagliflozin | 1 | -1.52 | NA | NA | NA |
| CDK inhibitor | aloisine | 1 | -1.51 | NA | NA | NA |
| CFTR channel agonist | aloisine | 1 | -1.51 | NA | NA | NA |
| GSK inhibitor | aloisine | 1 | -1.51 | NA | NA | NA |
| Glucocorticoid receptor antagonist | mifepristone | 1 | -1.51 | NA | NA | NA |
| Progesterone receptor antagonist | mifepristone | 1 | -1.51 | NA | NA | NA |
| Topoisomerase inhibitor | irinotecan | 1 | -1.51 | NA | NA | NA |
| ACE inhibitor | rescinnamine | 1 | -1.49 | NA | NA | NA |
| Antihypertensive | aliskiren | 1 | -1.48 | NA | NA | NA |
| Peptidase inhibitor | aliskiren | 1 | -1.48 | NA | NA | NA |
| Protease inhibitor | aliskiren | 1 | -1.48 | NA | NA | NA |
| Renin inhibitor | aliskiren | 1 | -1.48 | NA | NA | NA |
| Rho associated kinase inhibitor | fasudil | 1 | -1.48 | NA | NA | NA |
| Retinoid receptor agonist | palovarotene | 1 | -1.48 | NA | NA | NA |
| Calcitonin antagonist | telcagepant | 1 | -1.47 | NA | NA | NA |
| Angiogenesis inhibitor | tranilast | 1 | -1.47 | NA | NA | NA |
| Voltage-gated sodium channel blocker | fomocaine | 1 | -1.46 | NA | NA | NA |
| Farnesyltransferase inhibitor | geranylgeraniol | 1 | -1.46 | NA | NA | NA |
| DNA methyltransferase inhibitor | azacitidine | 1 | -1.45 | NA | NA | NA |
| Mucus protecting agent | mepazine | 1 | -1.45 | NA | NA | NA |
| Cholinergic receptor agonist | carbachol | 1 | -1.44 | NA | NA | NA |
| Dipeptidyl peptidase inhibitor | linagliptin | 1 | -1.44 | NA | NA | NA |

| | | | | | | |
|---|---|---|---|---|---|---|
| Thymidylate synthase inhibitor | fluorouracil | 1 | 1.95 | NA | NA | NA |
| LXR agonist | epoxycholesterol | 1 | 1.96 | NA | NA | NA |
